# Genomic basis of developmental defects of enamel and sex-specific effects

**DOI:** 10.64898/2026.07.06.26355672

**Authors:** P Shrestha, M Graff, Y Gu, Y Wang, HS Ahn, KN Nguyen, A Khanna, CL Avery, HM Highland, J Ginnis, MA Simancas-Pallares, AG Ferreira Zandoná, RN Alotaibi, DY Lin, JS Preisser, GD Slade, ML Marazita, KE North, K Divaris

## Abstract

We conducted a multi-ancestry genome-wide association study (GWAS) of developmental defects of enamel (DDE) in the primary dentition among 6,061 U.S. preschool-aged children (3–5 years). We investigated four DDE phenotypes (demarcated opacities, diffuse opacities, hypoplastic defects, and a combined DDE trait) leveraging main-effect models, joint gene-sex interaction testing (2df), and sex-stratified analyses. SNP-based heritability for the combined DDE trait was estimated at 20%, with concordance analyses robustly supporting a genetic etiology. We identified 39 unique genome-wide significant loci (*P*<5×10□□), with five surpassing a study-wide Bonferroni-corrected statistical significance criterion (*P*<1.25×10□□), including *Y RNA* and *ALDH1A1*. The main-effect GWAS identified 20 loci, including *HBS1L* and *MYB*, genes regulating hematopoiesis with plausible roles in amelogenesis. Joint test and sex-stratified analyses revealed 19 additional loci, including *ALDH1A1*, *TENM2*, and *DLGAP2*, demonstrating sex-specific heterogeneity. Nineteen loci exhibited sex-specific differences after Bonferroni correction (P<2x10^-3^), including genes involved in retinoic acid signaling (*ALDH1A1*), odontogenesis (*TENM2*), and neurodevelopment (*DLGAP2*, *CDH10*). Pathway enrichment highlighted ectodermal and synapse organization networks, suggesting shared etiological mechanisms between DDE and systemic conditions like neurofibromatosis and autism spectrum disorder. Notably, no locus generalized in an external GWAS of permanent dentition DDE, underscoring fundamental biological differences in the genetic architectures governing primary versus permanent enamel formation. Crucially, a comprehensive cross-trait pleiotropy lookup against early childhood caries (ECC) revealed no shared genetic architecture, supporting the notion that the established clinical and epidemiological association between DDE and ECC is likely driven by structural defects increasing caries lesion susceptibility rather than genetic pleiotropy. By integrating gene-sex interaction testing, this study offers novel insights into the complex, sexually dimorphic genetic etiology of DDE and augments the biological evidence base that can support the development of precision pediatric dentistry.

## Introduction

Developmental defects of enamel (DDE) are clinically manifest non-carious conditions resulting from perturbations during amelogenesis. Affecting 25% to 40% of the primary dentition globally, DDE encompass a heterogeneous group of conditions ranging from qualitative hypomineralization (e.g., demarcated and diffuse opacities) to quantitative hypoplastic defects (Seow et al. 2011; Robles et al. 2013). Beyond aesthetic concerns, these defects present a profound clinical burden (Jälevik and Klingberg 2002). Conditions such as molar-incisor hypomineralization (MIH) frequently cause severe hypersensitivity, rapid post-eruptive breakdown, and elevated caries risk. Consequently, DDE exert a substantial psychosocial and economic toll (Marshman and Rodd 2015), underscoring the urgency of elucidating their complex etiology.

While environmental factors such as early childhood illnesses and nutritional deficiencies are established as modifiers of enamel formation, amelogenesis is fundamentally governed by a highly conserved genetic program. Human candidate gene studies have reported associations between SNPs in enamel forming genes (Gerreth et al. 2018) and DDE. Moreover, animal models have demonstrated that disruptions in core enamel matrix genes (e.g., *ENAM*, *AMTN*, *KLK4*) result in characteristic enamel malformations (Hu et al. 2008; Núñez et al. 2016).

Furthermore, because the enamel organ shares an ectodermal embryonic origin with the central nervous system, and tooth mineralization overlaps with systemic developmental windows, enamel defects frequently exhibit pleiotropy with broader systemic conditions, including neurodevelopmental and hematopoietic disorders

Despite these insights, our understanding of the polygenic architecture of DDE, particularly in the primary dentition, remains incomplete. To-date, genome-wide association studies (GWAS) have primarily evaluated DDEs in permanent teeth and older cohorts (Kühnisch et al. 2014) (Alotaibi et al. 2022). Crucially, recent GWAS of complex dental phenotypes have highlighted the critical need to consider sex as a biological variable (Freitag-Wolf et al. 2021; Shrestha et al. 2025), as well as sex-based prevalence differences in DDE (Lunardelli et al. 2024). To address this knowledge gap and to add to the knowledge base of DDE genomic susceptibility, we sought to systematically identify genetic risk loci associated with DDE using a GWAS that leverages gene-sex interactions within a multi-ancestry population-based cohort of preschool-aged children.

## Methods

### Study design and population

The analysis sample comprised 6,061 participants with clinical DDE data from the ZOE 2.0 study, a community-based, multi-ethnic genetic-epidemiologic study of early childhood oral health (Divaris et al. 2020). Participants were 3–5-year-olds sampled from public preschools (Head Start) in North Carolina, United States, between August 2016 and February 2019. Details on study design, recruitment, exclusions, and the genotyped population are presented in the Appendix (**Appendix Figure 1**).

### Phenotype ascertainment

Trained clinical examiners used the modified DDE index to ascertain developmental defects of the enamel at the tooth level (Ginnis et al. 2019). Specifically, the facial/buccal surface of each was assessed with respect to the presence or absence of demarcated opacities (*dema*), diffuse opacities (*diff*), and hypoplastic defects (*hypo*). Based on these assessments, quantitative (count, with range=0-20) and binary (presence versus absence) traits were defined at the person level, including a combined DDE trait encompassing ‘any DDE’ (**Appendix Figure 2** and **Appendix Table 1**). To rigorously account for the right-skewness and zero-inflation inherent to count data (**Appendix Figure 3**), traits were regressed on age, sex, and race/ethnicity utilizing zero-inflated negative binomial models. The resulting Pearson residuals were carried forward to association testing.

### Genotyping, trait heritability, and concordance

Among the 6,262 genotyped saliva samples, 6,144 met strict quality control criteria. High-density genotyping and subsequent imputation via the Trans-Omics for Precision Medicine (TOPMed) server yielded ∼14.3 million high-quality single nucleotide polymorphisms (SNPs) (MAF >1%, R² >0.3). Details on quality control have been reported previously (Shrestha et al. 2025). The heritable variance (*h*^2^) of the DDE traits attributable to human genome was estimated among 5,580 unrelated participants using Genome-wide Complex Trait Analysis (GCTA) using high-quality imputed SNPs (R^2^>0.7); excluding SNPs with MAF<5%; and adjusting for age, sex, eight ancestry principal components and self-reported race/ethnicity (Yang et al. 2013). To further evaluate DDE’s genetic underpinning, we additionally estimated trait concordance among 682 related pairs utilizing Cohen’s *kappa* and intraclass correlation coefficients (ICC).

### Genome-wide association analyses

To investigate the genetic basis of DDE and interrogate potential sex dimorphism, we employed SAIGE (Zhou et al. 2018) to efficiently account for sample relatedness and case-control imbalance in a three-tiered approach (**Figure 1**): a) **Approach 1** (**main effects**). Discovery GWAS (n=6,061) using linear mixed models for the quantitative traits (utilizing the aforementioned zero-inflated negative binomial Pearson residuals) and logistic mixed models for the binary traits. b) **Approach 2 (joint gene-sex testing)**. Joint 2-degree-of-freedom (2df) testing to simultaneously evaluate genetic main effects and gene-sex (GxS) interactions, a method optimizing power for discovering loci with heterogeneous effects between sexes (Kraft et al. 2007; Manning et al. 2011). C) **Approach 3** (**sex-stratified models**). The sample was split into male and female strata, to fit separate sex-stratified main-effects models. We subsequently calculated the P-value for difference (P*_diff_*) between the stratum-specific beta coefficients to formally screen for GxS interactions.

**Figure 1:**
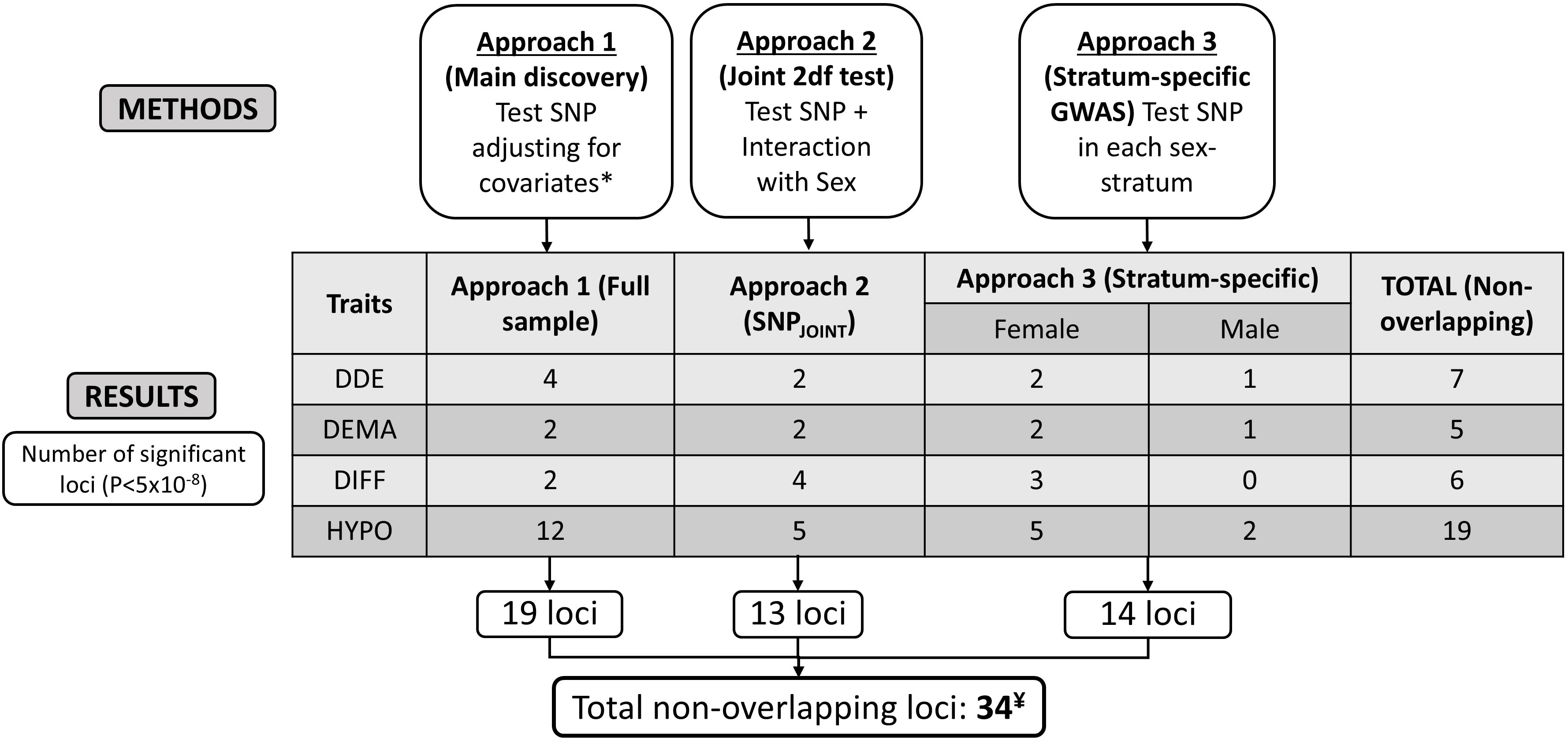
Summary of study design and results for primary count traits. Approach 1 conducted a genome-wide association study (GWAS) in the entire sample. Approach 2 conducted a joint 2-degree-of-freedom test to test the main and interaction effects jointly accounting for sex. Approach 3 conducted sex-specific GWAS of the developmental defects of enamel primary quantitative traits. The approaches have been detailed in the Methods section. The total number of loci is less than the sum of the boxes owing to the identification of the same loci in different traits or analyses. *Covariates include age, sex, race/ethnicity, and the first 8 principal components of ancestry. ¥ Analysis of secondary binary traits identified further 5 independent genomic risk loci. Abbreviations: DDE: Developmental Defects of Enamel, DEMA: Demarcated Opacities, DIFF: Diffuse Opacities, HYPO: Hypoplastic defects, SNP: Single Nucleotide Polymorphism, 2df: 2-degree-of-freedom, GWAS: Genome-wide association study, P: p-value.

EasyStrata was used to carry out QC, generate Manhattan and quantile-quantile (Q-Q) plots, and conduct the 1df (sex-specific), 2df (P*_joint_*) and the P*_diff_* tests (Winkler et al. 2015)

### Statistical testing, annotation, and generalization

We applied a conventional genome-wide significance threshold (*P*<5×10□□) to detect novel loci, and a stringent study-wide Bonferroni-corrected genome-wide statistical significance threshold (*P*<1.25×10□□) to account for multiple testing (i.e., total 40 models emanating from 8 phenotypes interrogated in 5 models). We excluded SNPs with effective allele sample size (effN) <100 from reporting genome-wide significant association results. We utilized FUMA GWAS for functional annotation (Watanabe et al. 2017), alongside MAGMA v1.6 for gene-centric and pathway enrichment analyses.

To evaluate the generalization of our findings, we examined the directional consistency and Bonferroni-corrected statistical significance (*P*<1.3×10□³) of our top SNPs against summary statistics from external cohorts: a multi-ethnic GWAS of enamel hypoplasia (Alotaibi et al. 2022; n=7,159), a childhood caries meta-analysis (Haworth et al. 2018; n=19,000), and an adult caries meta-analysis (Shungin et al. 2019; n=487,000). Finally, to investigate whether the established clinical association between DDE and early childhood caries (ECC) is driven by shared genetics, we conducted a comprehensive cross-trait pleiotropy look-up. We examined 939 trait-variant associations against primary (cavitated lesions; ICDAS≥3) and secondary (clinically-manifest; ICDAS≥1) ECC phenotypes using summary statistics from a recent multi-ancestry GWAS of ECC conducted within the same ZOE 2.0 cohort (Shrestha et al. 2025; n=6,103). This look-up systematically evaluated both main effects and gene-environment interactions (i.e., interactions with fluoride and sugary beverage consumption frequency) to estimate the proportion of loci reaching nominal or genome-wide significance.

## Results

The study’s analytical sample (n=6,061) included 50% females with a mean age of 53 months. Approximately half were non-Hispanic African Americans (48%) with 20% Hispanic Americans, 18% Non-Hispanic Whites, and 2% American Indian/Alaskan Natives (**Appendix Table 3**). To evaluate the genetic underpinning of DDE, we estimated SNP-based heritability (*h*²) and familial concordance. Common SNPs explained 20% of the phenotypic variance for the combined DDE quantitative trait (*h*²=0.20, se=0.06, P=7.9×10□□). Among distinct subtypes, heritability was highest for diffuse opacities (*h*²=0.16), with estimates for other traits <10% (**Appendix Table 4**). As expected for heritable traits, familial concordance increased with the degree of relatedness. Concordance for the combined DDE quantitative trait was highest among monozygotic twins (ICC=0.43, 95% CI: 0.15–0.65), intermediate for first-degree relatives (ICC=0.41, 95% CI: 0.30–0.50), and lowest for second- and third-degree relatives (ICC=0.19, 95% CI: 0.09–0.28) **(Appendix Table 4)**.

Across the three analytical approaches, genomic inflation factors (λ_GC_) ranged between 0.93 and 1.01, indicating good control of population stratification (**Appendix Table 5; Appendix Figure 4**). We identified 39 unique genome-wide significant loci (*P*<5×10□□) across all models, with five (*Y RNA, RP11-157E21.2, AC007950.1, KRTAP7-1, ALDH1A1*) surpassing a stringent study-wide Bonferroni-corrected threshold (*P*<1.25×10□□) (**Figure 1**). The main-effect GWAS identified 20 unique loci reaching genome-wide significance (**Table 1**, **Figure 2**). Nineteen of these loci were associated with quantitative traits, representing 20 distinct trait-locus associations (i.e., 4 for any DDE, 2 for demarcated opacities, 2 for diffuse opacities, and 12 for hypoplastic defects) (**Table 1**, **Table 2**). Notably, the *Y RNA* locus provided a strong association signal for both diffuse opacities and overall DDE. Moreover, *RP1-281H8.3* was significantly associated with the diffuse opacities binary trait.

**Table 1.** Novel loci emerging from the genome-wide association analyses conducted for developmental defects of enamel in the full sample of the ZOE 2.0 study (Approach 1)

| Locus | Lead SNP (rsid) | EA/OA | EAF | effN | b | SE | P | Imputed |
| --- | --- | --- | --- | --- | --- | --- | --- | --- |
| Quantitative traits (count of affected teeth) |  |  |  |  |  |  |  |  |
| Developmental defects of enamel |  |  |  |  |  |  |  |  |
| *Y_RNA*<sup>*</sup> | rs143210152 | G/C | 0.02 | 174 | 0.45 | 0.08 | 4.4E-09 | Yes |
| *DEK* | rs567243779 | A/C | 0.06 | 619 | 0.23 | 0.04 | 8.1E-09 | Yes |
| *RP4-724E13.2* | rs150075871 | T/C | 0.01 | 119 | 0.53 | 0.09 | 2.0E-08 | Yes |
| *HBS1L* | rs116989668 | A/G | 0.01 | 120 | 0.52 | 0.10 | 4.0E-08 | Yes |
| Demarcated opacities |  |  |  |  |  |  |  |  |
| *SLCO4A1* | rs75880235 | T/C | 0.01 | 132 | 0.51 | 0.09 | 1.9E-08 | No |
| *C21orf54* | rs895438538 | CA/C | 0.03 | 298 | 0.33 | 0.06 | 4.2E-08 | Yes |
| Diffuse opacities |  |  |  |  |  |  |  |  |
| *Y_RNA*<sup>‡</sup> | rs77669438 | A/G | 0.01 | 142 | 0.56 | 0.09 | **9.1E-11** | Yes |
| *CTA-21C21.1*<sup>*</sup> | rs1658697761 | CA/C | 0.41 | 2914 | 0.11 | 0.02 | 2.0E-09 | Yes |
| Hypoplastic defects |  |  |  |  |  |  |  |  |
| *RP11-157E21.2*<sup>*</sup> | rs192067416 | G/A | 0.02 | 205 | 0.46 | 0.07 | **2.4E-10** | Yes |
| *AC007950.1*<sup>*</sup> | rs79505735 | C/T | 0.02 | 189 | 0.46 | 0.07 | **5.1E-10** | Yes |
| *RP1-167G20.1*<sup>*</sup> | rs149507176 | C/T | 0.01 | 143 | 0.52 | 0.09 | 1.5E-09 | Yes |
| *ADAMTSL1* | rs115373518 | G/A | 0.03 | 342 | 0.32 | 0.06 | 7.5E-09 | Yes |
| *RAVER2* | chr1:64830234 | T/TG | 0.01 | 131 | 0.50 | 0.09 | 7.6E-09 | Yes |
| *FHIT* | rs73835238 | G/A | 0.01 | 129 | 0.52 | 0.09 | 9.1E-09 | No |
| *UBE2G2* | rs115617269 | A/G | 0.02 | 172 | 0.43 | 0.08 | 2.0E-08 | Yes |
| *MPV17L2* | rs79028494 | A/G | 0.04 | 408 | 0.28 | 0.05 | 2.9E-08 | Yes |
| *PPAPDC3* | rs75061854 | T/C | 0.02 | 196 | 0.41 | 0.07 | 2.9E-08 | Yes |
| *CLUL1* | rs114989924 | G/A | 0.02 | 232 | 0.37 | 0.07 | 3.6E-08 | Yes |
| *MAML2* | rs150489138 | A/G | 0.01 | 147 | 0.45 | 0.08 | 4.3E-08 | Yes |
| *MTHFD1L* | rs11963290 | T/C | 0.11 | 1160 | 0.17 | 0.03 | 4.8E-08 | Yes |
| Binary traits (case status) |  |  |  |  |  |  |  |  |
| Diffuse opacities |  |  |  |  |  |  |  |  |
| *RP1-281H8.3* | rs184135739 | C/T | 0.01 | 153 | 0.98 | 0.17 | 3.0E-09 | Yes |

**Boldface** signifies meeting the stringent study-wide Bonferroni-corrected genome-wide statistical significance threshold ( $P < 1.25 \times 10^{-8}$ )
<sup>\*</sup> Denotes SNP is also genome-wide statistically significant for the joint test (main genetic effect + gene x environment effect)
<sup>‡</sup> Denotes SNP is genome-wide significant for more than one trait (e.g., diffuse opacities and overall DDE)
Abbreviations: Chr - chromosome; EA: Effect allele; effN: effective allele count; OA: Other allele; EAF: effect allele frequency; SNP - Single nucleotide polymorphism

**Figure 2:**
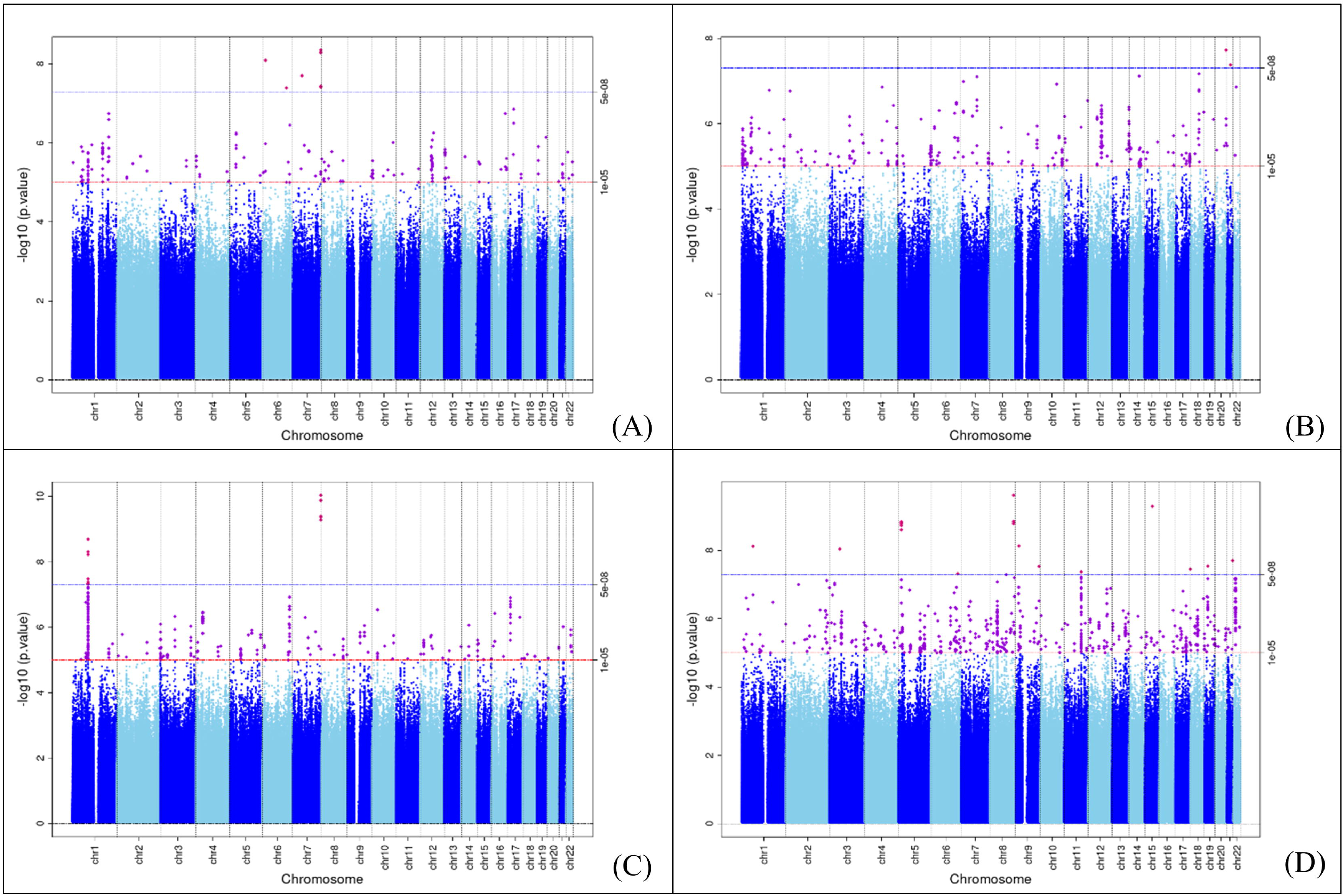
Manhattan plots for the primary quantitative traits with genome-wide statistically significant loci (Approach 1). A) DDE, B) Demarcated opacities, C) Diffuse Opacities, D) Hypoplastic defects

**Table 2.**
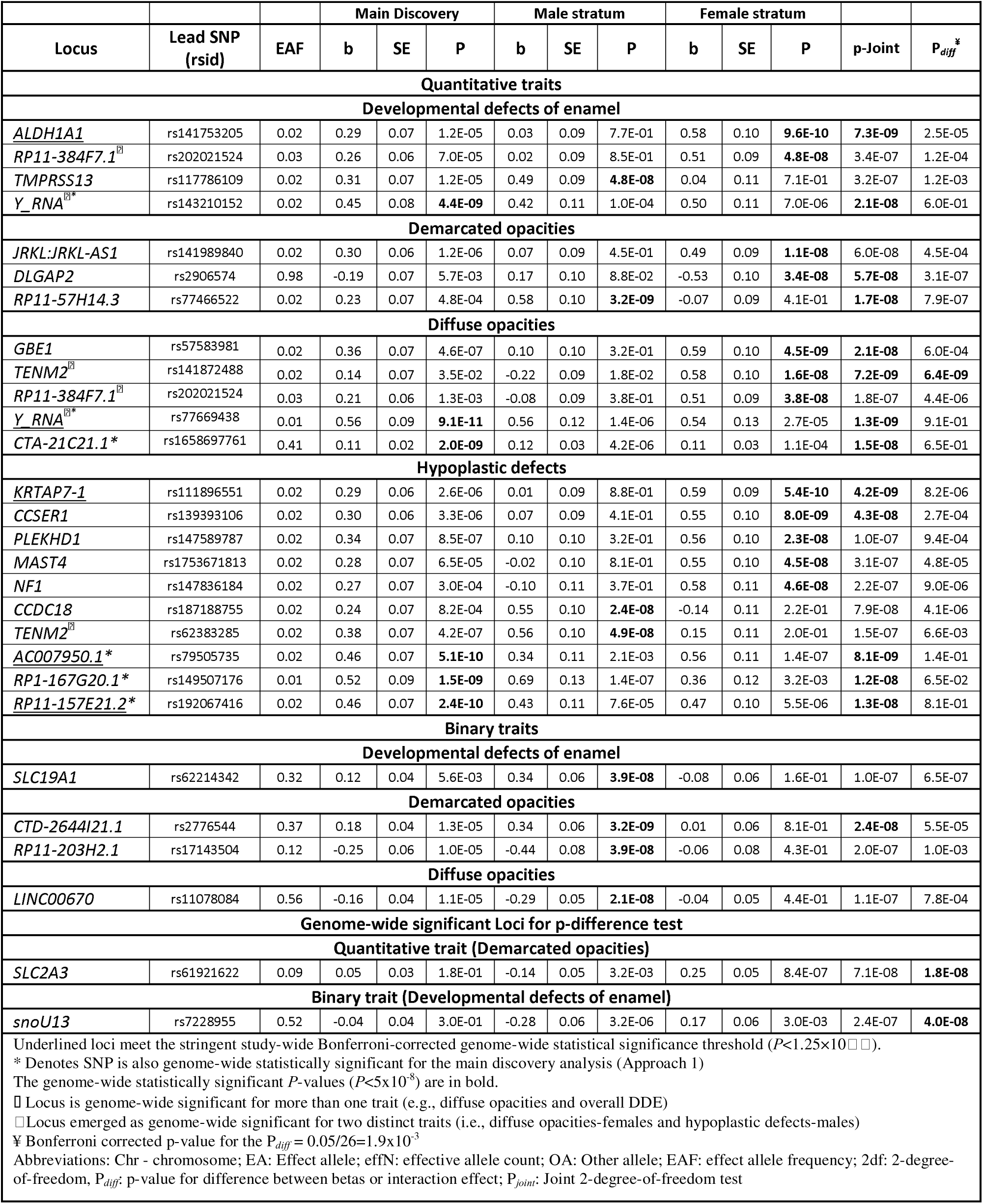
Summary of association results for the novel genetic risk loci identified in Approaches 2 and 3.

The joint 2-degree-of-freedom tests (Approach 2) and sex-stratified models (Approach 3) provided additional biological sex-informed insights, nominating 19 additional unique loci (e.g., *ALDH1A1*, *TENM2*, and *DLGAP2*). Sex-stratified analyses illuminated signals driven by male or female strata; e.g., *RP11-384F7.1* reached genome-wide significance exclusively in the female stratum for both the combined DDE and diffuse opacities traits (**Table 2**). Nineteen loci exhibited statistically different effect sizes between males and females using a Bonferroni correction (*P_diff_*<2×10□³) (**Figure 3**), underscoring a sexual dimorphism in DDE genetics.

**Figure 3:**
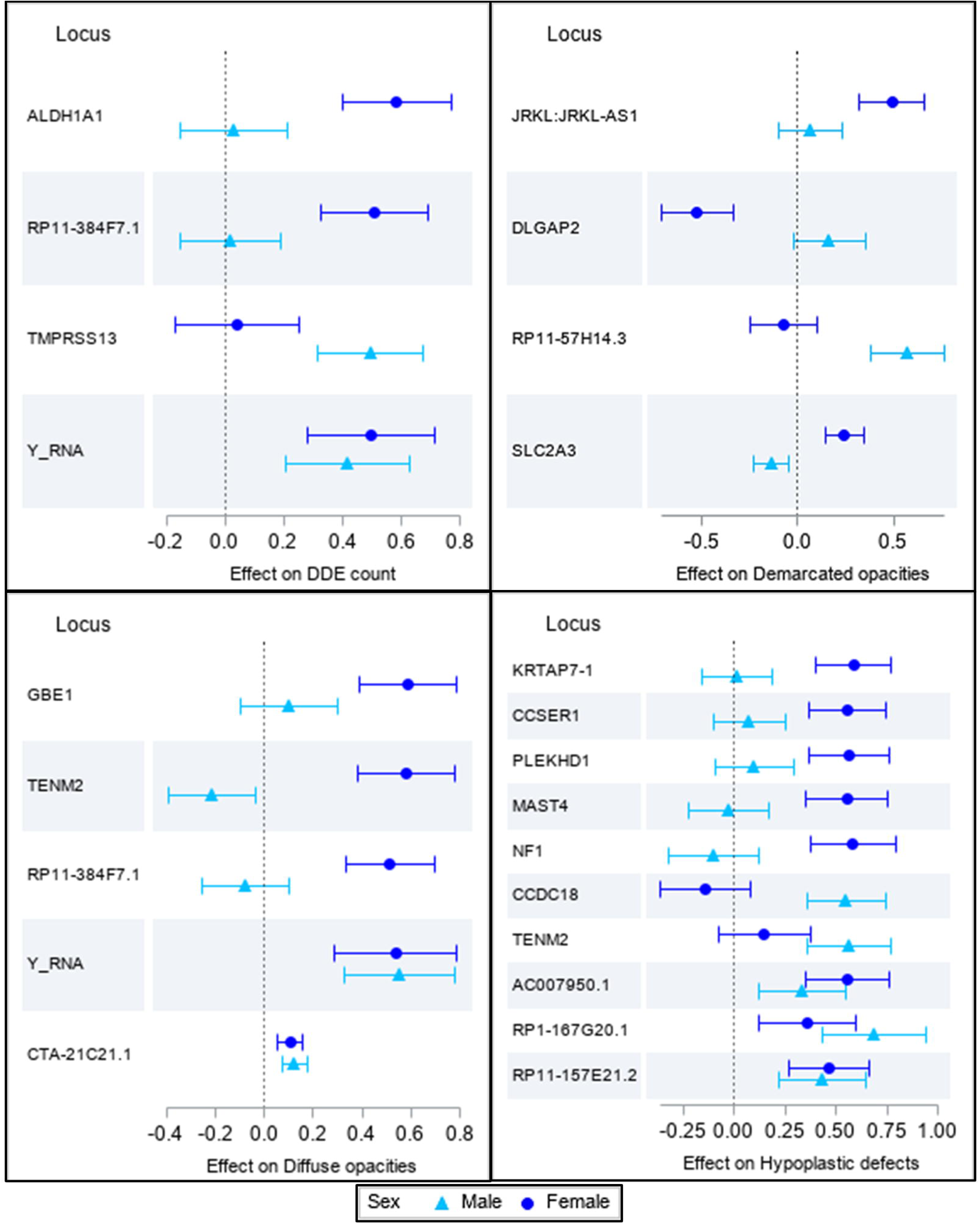
Forest plots demonstrating heterogeneity of genetic effect in different sexes for the 4 primary traits. (A) Developmental defects of enamel count, (B) Demarcated opacities, (C) Diffuse opacities, (D) Hypoplastic defects. The triangles and circles represent effect allele estimates of association and the error bars represent the corresponding 95% confidence intervals.

Strikingly, three loci demonstrated significant sex-specific heterogeneity at a strict genome-wide association threshold: *SLC2A3* (*P_diff_*_=_1.8×10□□), *TENM2* (*P_diff_*=6.4×10□□), and *snoU13* (P_diff=_4.0×10□□).

To prioritize variants, we annotated significant loci using CADD and RegulomeDB (RDB) scores (**Appendix Tables 6 and 7)** and tissue-specific gene expression (**Research Data Table 1)**. The biological roles of nearby (∼250Kb) protein-coding genes are detailed in **Research Data Tables 2 and 3**. We identified multiple loci harboring highly deleterious, non-coding regulatory variants that serve as prime candidates for future *in vitro* functional validation assays. For diffuse opacities, variants in high LD with *CTA-21C21.1* had high CADD scores (e.g., rs566606 CADD: 21.4). Similarly, for hypoplastic defects, potentially functional regulatory variants were identified at *CLUL1* (rs114989924; CADD: 13.0) and *MAML2* (rs142941402; CADD: 17.3).

Gene-based testing via MAGMA mapped SNPs to 18,273 protein-coding genes and identified *CDH10* as genome-wide significant. *CDH10* encodes a cadherin protein, plays a crucial role in synaptic adhesion, axon guidance, and maintaining tissue structure, and has been previously associated with autism spectrum disorder (ASD) (Redies et al. 2012). Further pathway enrichment revealed links to neurodevelopmental conditions, with the *DLGAP2* locus (synapse organization) emerging for demarcated opacities, and the *NF1* locus (neurofibromatosis type-1) emerging for hypoplastic defects. Gene-set analyses identified three sets reaching Bonferroni-corrected significance for hypoplastic defects “GO_bp: go_elastic_fiber_assembly” and “Curated_gene_sets: valk_aml_with_t_8_21_translocation” from Approach 1, and “GO_cc: go_stereocilium_membrane”, and enrichment of the prioritized genes from Approach 1 for hypoplastic defects within the facial emotion recognition gene set (**Appendix Table 10**).

No signal generalized to the external cohorts with GWAS of enamel hypoplasia and dental caries (**Appendix Tables 11–14**) using strict multiple-testing-corrected criteria, likely reflecting phenotypic and demographic differences between the cohorts. However, one locus, *SLC19A1*, was nominally significant and directionally consistent with a previous GLIDE meta-analysis of primary teeth. While exact variants did not broadly generalize, a regional examination (±120 Kb) of dental caries GWAS data identified strong, co-localizing signals at two loci on chromosome 15 (*AC007950.1*, GLIDE adult caries, *P*=1.3x10^-26^) and chromosome 6 (*DEK*, clinical decay, *P*=1.8x10^-7^) (**Appendix Figure 6**). Cross-trait look-ups were carried out to illustrate the contribution of GWAS signals across multiple distinct DDE clinical subtypes and the overall DDE trait (**Appendix Table 15**).

To evaluate possibly shared genetic susceptibility underlying the established DDE-ECC clinical association, we conducted a cross-trait pleiotropy look-ups evaluating 939 DDE trait-SNP associations against ECC phenotypes (Shrestha et al. 2025). Examining main effects and gene-environment interactions, we found no greater-than-chance associations, with only 40 SNP look-ups (4.3%) reaching nominal significance (*P*<0.05) and none reaching genome-wide significance, as expected under the null hypothesis of no association. This finding is supportive of the notion that a DDE-ECC clinical and epidemiologic association is most likely structural (i.e., tooth surface susceptibility) rather than driven by pleiotropy (i.e., shared genetics).

## Discussion

To further our understanding of the polygenic underpinning of developmental defects of the enamel, we conducted the largest multi-ancestry GWAS of primary dentition DDE. Our findings suggest a considerable genetic basis, with common SNPs explaining approximately 20% of the phenotypic variance of DDE. By integrating main-effect models with joint gene-sex interaction testing and sex-stratified analyses, we identified a total of 39 unique genome-wide significant risk loci. Some of the newly identified loci (e.g., in *CTA-21C21.1* and *MAML2*) harbor SNPs with exceptionally high CADD scores, representing highly deleterious, non-coding regulatory variants, which we propose as prime candidates for future functional validation assays. Crucially, the results support shared biological mechanisms with odontogenesis, hematopoiesis, and neurodevelopment, a profound sexual dimorphism in DDE susceptibility, and demonstrate that the established clinical association between DDE and childhood caries is likely driven by structural enamel defects rather than genetic pleiotropy.

The main discovery analysis in the analysis sample (Approach 1) identified 19 loci with several harboring genes with highly plausible biological roles in amelogenesis and systemic health. For the combined DDE trait, the *HBS1L* locus emerged as highly significant. *HBS1L* encodes a GTP-binding elongation factor associated with sickle cell disease (Safran et al. 2021), aligning with epidemiological reports of a higher DDE prevalence among patients with sickle cell disease (Lopes et al. 2018). Located ∼170 Kb from the lead variant near *HBS1L* is *MYB*, a transcription regulator that is strongly expressed in odontoblasts and ameloblasts during embryonic tooth development, with a well-documented role in tooth mineralization (Matalová et al. 2011).

Among the loci identified for hypoplastic defects, *FHIT* is a tumor suppressor previously associated with odontogenic tumors and non-syndromic cleft lip/palate with hypodontia (Malčić et al. 2008; Ghazali et al. 2021). We also identified signals near *TSPEAR*, which regulates the Notch signaling pathway; the latter is a critical driver of ameloblast derivation from the ectoderm and overall enamel development (Cai et al. 2011; Peled et al. 2016). Furthermore, the diffuse opacities-associated *LATS1* encodes a kinase regulating the Hippo signaling pathway, which is essential for the development of the enamel organ (Wang et al. 2022). Additional loci included *IL10RB* (near *C21orf54*), previously linked to MIH (Elzein et al. 2022), and *NTSR1* (near *SLCO4A1*), a neurotensin mediator associated with dental care-related anxiety (Zhou et al. 2022). For the secondary binary traits, *COL18A1* (near *SLC19A1*) and *DLK1* have been implicated in enamel matrix maturation and ameloblast proliferation, respectively (Väänänen et al. 2004; Qi et al. 2018).

Differences in genetic effects between sexes have been reported for several common-complex dental, oral, and craniofacial diseases including ECC and early-onset periodontitis (Freitag-Wolf et al. 2021; Shrestha et al. 2025). By leveraging joint 2-degree-of-freedom tests and sex-stratified GWAS, we identified 19 additional loci marked by sex-specific heterogeneity. For example, *ALDH1A1*, which encodes an enzyme responsible for retinoic acid synthesis, strongly emerged in female-specific and joint models. *ALDH1A1* has been shown to be downregulated during the mid-late maturation stage of enamel formation relative to the secretory stage, highlighting a sex-mediated biological effect in amelogenesis (Lacruz et al. 2012). Similarly, *TENM2* exhibited distinct risk profiles between males and females and has been previously shown to be associated with periodontal pathogen colonization and moderate periodontitis (Divaris et al. 2012); (Hong et al. 2015). The finding of 19 loci exhibiting statistically different effect sizes between males and females (e.g., *SLC2A3*, *snoU13*, and *TENM2* reaching genome-wide significance for interaction) underscores the notion that analyzing sexes in aggregate may mask potentially informative genetic heterogeneity and supports the necessity of treating sex as a biological variable in dental genomics (Shrestha et al. 2025).

The enamel organ and the central nervous system share an ectodermal embryonic origin, and it is thus unsurprising that our results demonstrate a genetic overlap of DDE with neurodevelopmental disorders, with gene-based association testing and pathway enrichment analyses strongly supporting this overlap. For example, we identified *CDH10* (diffuse opacities) and *DLGAP2* (demarcated opacities), both of which play crucial roles in synapse organization and are established candidate genes for ASD (Redies et al. 2012) (Rasmussen et al. 2017). This finding provides a genomic basis for clinical observations showing a higher incidence of enamel defects in children with intellectual disabilities and ASD (Modrić et al. 2016). Similarly, we identified the *NF1* locus for hypoplastic defects; neurofibromatosis type-1 is a condition known to present with an elevated prevalence of DDE (Santoro et al. 2020). While additional research is required to substantiate and elucidate these associations, these shared genetic pathways support the notion that disruptions during overlapping developmental windows may manifest pleiotropically across multiple ectoderm-derived tissues.

Clinically, individuals and teeth with DDE are known to be at a high risk of developing ECC (Caufield et al. 2012; AAPD 2025). As demonstrated by our cross-trait pleiotropy look-up against ECC phenotypes (Shrestha et al. 2025), this established clinical association is almost certainly driven by structural vulnerability rather than shared genetics. Because DDE constitutes altered enamel formation primarily in utero or during early infancy, affected teeth erupt with pre-existing structural deficits. This pre-existing structural susceptibility rather than a shared genetic susceptibility, is what is likely driving ECC susceptibility when faced with a cariogenic, post-eruptive, dysbiotic biofilms.

None of our 39 genome-wide significant loci generalized in an external multi-ethnic GWAS of enamel hypoplasia (Alotaibi et al. 2022). The lack of replication is likely a consequence of a fundamental difference in phenotypes and underlying biology: Alotaibi et al. investigated permanent teeth in an older cohort (7–82 years), whereas the present report exclusively focused on primary teeth of 3-5-year-olds. Because primary enamel forms *in utero* under maternal physiological influences, whereas permanent enamel mineralizes post-natally, their genetic underpinnings likely differ significantly. We did, however, observe nominal significance and directional consistency for *SLC19A1* in a GLIDE meta-analysis of primary teeth, as well as a nominal overlap for rs13058467 near *SCUBE1*, a suggestive MIH locus previously reported by (Kühnisch et al. 2014) (**Appendix Table 16**).

Strengths of this study include its multi-ancestry, community-based design, which improves statistical power, refines signals across populations, and enhances public health relevance (Li and Keating 2014). The rigorous modeling of zero-inflated quantitative traits and the large sample size represent substantial methodological improvements over prior GWAS of DDE. The innovative incorporation of joint 2df testing maximized our power to uncover latent heritability driven by gene-sex interactions (Manning et al. 2011). However, several limitations must be acknowledged. The scarcity of comparable primary dentition DDE GWAS data prevented validation in an independent, age-matched replication cohort. Additionally, because severe forms of DDE (e.g., widespread and severe hypoplastic defects of the enamel) can rapidly undergo post-eruptive breakdown and serve as the basis of extensive caries lesion development, it is conceivable that some DDE phenotypes were masked by ECC during clinical examinations, leading to an under-estimation of the phenotype. Finally, some of our identified loci are driven by less common variants (MAF between 1–5%), increasing the potential for false positives.

While we identified non-coding regulatory variants with exceptionally high CADD scores indicative of deleterious regulatory effects (e.g., in *CTA-21C21.1*, *CLUL1*, and *MAML2*), functional validation via *in vitro* and *in vivo* mechanistic experiments is warranted to confirm their roles in amelogenesis.

In conclusion, this study reports valuable evidence on the genomic architecture of developmental defects of enamel in the primary dentition. It highlights 39 novel candidate loci, addressing a knowledge gap in the genomic basis of amelogenesis, and offers several plausible links between systemic neurodevelopmental and hematopoietic pathways and ECC. At the same time, our data support the notion that the clinical and epidemiological link between DDE and ECC is likely driven by structural enamel defects rather than genetic pleiotropy. Importantly, the profound sex-specific genetic effects reported in this study underscore the importance of treating sex as a biological variable in dental genomics investigations. Upon further functional validation, these discoveries have the potential to enhance our biological understanding of enamel development, augment ECC risk assessment tools, and advance future applications in precision pediatric oral health.

## Supporting information

Main Figures

Research Data

## Data Availability

The genotype and phenotype datasets from ZOE 2.0 are publicly available through phs002232.v1.p1 “TOPDECC-Trans-omics for Precision Dentistry and Early Childhood Caries: Genome-Wide Genotyping (CIDR) and Microbiome in the ZOE 2.0 Study”. Genomic summary results of the GLID-adults study are available at: https://data.bris.ac.uk/data/dataset/2j2rqgzedxlq02oqbb4vmycnc2 and GLIDE-children at: https://data.bris.ac.uk/data/dataset/108de7fd6f8ff188ebb9ac07fe77bfb5.

## ACKNOWLEDGEMENTS

This work was supported by research grants from the National Institutes of Health: National Institute for Dental and Craniofacial Research U01DE025046 (K.D., A.G.F.Z, D.Y.L, J.S.P, G.D.S., K.E.N) and National Human Genome Research Institute X01HG010871 (K.D.). K.D. and P.S. are supported by the National Institute of Dental and Craniofacial Research (NIDCR), National Institutes of Health (NIH), under award R01DE034410. The content is solely the responsibility of the authors and does not necessarily represent the official views of the funders.

The authors thank CIDR investigators and staff at Johns Hopkins University for carrying out genotyping and imputation for the project with support from a resource-allocation grant NIH/NIHGR X01-HG010871; Dr. Patricia V. Basta and her team at the UNC-Chapel Hill Biospecimen Processing facility for the accessioning, storage, and disbursement of the saliva and extracted nucleic acid samples in the ZOE studies; all study participants and their families for their contributions.

## Author Contributions

P. Shrestha, contributed to data analyses and annotation, results interpretation, drafted and critically revised the paper. M. Graff, contributed to data analyses and annotation, results interpretation, and critically revised the paper. Y. Gu, contributed to data analyses and annotation, and critically revised the paper. Y. Wang, contributed to data analyses and annotation, and critically revised the paper. H.S. Ahn, contributed to data analyses and annotation, and critically revised the paper. K.N. Nguyen, contributed to data analyses and annotation, and critically revised the paper. A. Khanna, contributed to data analyses and annotation, and critically revised the paper. C.L. Avery, contributed to results interpretation, and critically revised the paper. H.M. Highland, contributed to data analyses and annotation, and critically revised the paper. J. Ginnis, contributed to data collection, and critically revised the paper. M.A. Simancas-Pallares, contributed to data collection, and critically revised the paper. A.G. Ferreira Zandoná, contributed to study design, and critically revised the paper. R.N. Alotaibi, contributed to data analyses and annotation, and critically revised the paper. D.Y. Lin, contributed to results interpretation, and critically revised the paper. J.S. Preisser, contributed to study design, results interpretation, and critically revised the paper. G.D. Slade, contributed to study design, and critically revised the paper. M.L. Marazita, contributed to results interpretation, and critically revised the paper. K.E. North, contributed to conceptualization, study design, supervision, results interpretation, and critically revised the paper. K. Divaris, contributed to conceptualization, study design, supervision, data collection, results interpretation, and critically revised the paper. K. Divaris and K.E. North contributed equally. All authors gave their final approval and agreed to be accountable for all aspects of the work.

