## Appendix for "Genomic basis of developmental defects of enamel and sex-specific effects"

**APPENDIX NOTES**

**1. Description of different types of developmental defects of enamel**

### 1. **Hypoplastic defects**

Hypoplastic defects are characterized by a reduced thickness of the enamel, manifesting clinically as pits, grooves, or broader areas of missing enamel. Pathologically, these defects occur due to insults during the secretory phase of amelogenesis (Suckling 1989). Sarnat and Schour (Sarnat and Schour 1942) suggested that a narrower defect is formed due to a short-duration insult resulting from an acute systemic disease, while a wider defect is due to a longer-duration insult associated with a chronic disease or insult. However, Suckling noted that the insults producing a hypoplastic defect are generally of short duration, and the insult's severity fundamentally determines the extent of the lesion and the translucency of the affected enamel. Moreover, the specific etiology is not considered to be a major determining factor in the distinct presentation of these lesions (Suckling 1989).

### 2. **Demarcated opacities**

Demarcated opacities are characterized by an alteration in the translucency of the enamel presenting with normal thickness and a smooth surface. They exhibit a clear, distinct boundary (i.e., are well demarcated) with the adjacent normal enamel and can appear white, yellow, brown, or cream in color. According to experimental models by Suckling, these defects can occur due to an insult in either the secretory or maturation phases of amelogenesis (Suckling 1989). It is noted that the insults causing demarcated opacities are longer-lasting and less severe than those responsible for hypoplasia.

### 3. Diffuse opacities

Diffuse opacities present as an alteration in the translucency of the enamel, maintaining normal thickness and a smooth surface, but lacking a clear border with the adjacent normal enamel. They are typically white in color and present with a linear, patchy, or confluent distribution. Although originally hypothesized to occur due to an insult during the maturation phase, Suckling (Suckling 1989) demonstrated in an ovine model that the introduction of a low-dose of fluoride during the secretory phase presented with more severe diffuse opacities. A long duration of a low-grade insult has been heavily implicated in the pathogenesis of diffuse opacities. Although fluoride is a commonly reported modifier for diffuse opacities, other systemic factors including malnutrition, chronic illnesses, or high altitude, can replicate or exacerbate the effects of fluoride (Suckling 1989; Wong 2014).

**2. Methods**

**Study design and population**

ZOE 2.0 is a community-based, cross-sectional, multi-ethnic genetic epidemiologic study that enrolled 8,059 children ages 3-5 years from public preschools (Head Start) in North Carolina between 2016 and 2019. Details of the study design have been reported previously (Divaris et al. 2020). A flowchart depicting recruitment, enrollment, exclusions, and the genotyped population eligible for analysis (n=6,144) is presented in **Appendix Figure 1**. To account for high genetic relatedness, we excluded one sibling from each of the 41 monozygotic (MZ) twin pairs to produce the final analysis sample (n=6,103). Among these 6,103 individuals, clinical data for developmental defects of enamel (DDE) were missing for 42 participants, resulting in a final analytical sample of 6,061. The study was reviewed and approved by the University of North Carolina at Chapel Hill Office of Human Research Ethics Institutional Review Board on September 18, 2014 (IRB# 14-1992).

**Phenotypes**

Clinical examinations were carried out by trained examiner-recorder pairs (Ginnis et al. 2019). DDE ascertainment was performed on non-desiccated teeth, assessing the facial/buccal surfaces of all teeth present in the oral cavity. Tooth surface conditions were systematically assessed for the type and extent of DDE if the defect was ≥1 mm in diameter. We utilized Clarkson and O’Mullane’s modified DDE epidemiologic index to measure DDE at the tooth level (Clarkson and O’mullane’ 1989). For these analyses, we defined 8 phenotypes: quantitative (count) and binary (presence/absence) variables for demarcated opacities, diffuse opacities, hypoplastic defects, and a combined DDE trait encompassing all three (A**ppendix Figure 2** and **Appendix Table 1**). The 4 quantitative count variables served as primary traits, and the 4 binary variables served as secondary traits.

**Genotype data**

Saliva samples were collected using the DNA Genotek Oragene DNA-575 kit (DNA Genotek, Ottawa, Ontario, Canada) from 6,369 participants. DNA was extracted from the 6,262 samples meeting strict quality and quantity criteria, with a total of 6,144 study samples genotyped following quality control (QC) checks. High-density genotyping was carried out at the Center for Inherited Disorders Research (CIDR) at Johns Hopkins University, using the Infinium™ Global Diversity Array-8 v1.0 (Illumina, San Diego, CA, USA). CIDR technical failures, missing call rates (≥2%), >1 discordant call in 62 study duplicates, >1 Mendelian error, Hardy-Weinberg equilibrium deviation (*P* < 8×10⁻⁴), sex difference in allele frequency (≥0.2) or heterozygosity (>0.3) for autosomes/XY, and positional duplicates were utilized as rigorous QC metrics to filter SNPs, resulting in the release of 1,821,357 high-quality genotyped SNPs. Genotype imputation was carried out for 6,103 study participants via the Trans-Omics for Precision Medicine (TOPMed) imputation server. One sibling from each of the 41 MZ twin pairs was included in the imputation process. A total of 306,740,382 SNPs were imputed, informed by 1,142,635 genotyped SNPs overlapping the reference panel. Among these, 143,332,340 (46.7%) were non-monomorphic variants eligible for downstream processing. We further applied minor allele frequency (MAF) >0.01 and imputation quality score (*R²*) >0.30 filters, yielding 14,371,011 robust SNPs for the genome-wide association analyses (**Appendix Table 2**). Details regarding QC and methodological derivations (e.g., principal components and relatedness) have been reported previously (Shrestha et al. 2025).

**Heritability estimates**

The heritable variance (*h²*) of DDE traits attributable to common human genetic variation was estimated among 5,580 unrelated participants utilizing Genome-wide Complex Trait Analysis (GCTA) with high-quality imputed SNPs (*R²* > 0.7). SNPs with a MAF <5% were excluded. Models were adjusted for age, sex, eight ancestry principal components, and self-reported race/ethnicity (Yang et al. 2013). GCTA employs a two-step method: first, estimating a genetic relationship matrix (GRM) using all available SNPs for all individuals; second, carrying the GRM forward to a restricted maximum likelihood analysis to estimate the total proportion of phenotypic variance explained by all included SNPs.

**Statistical analyses**

*Modelling considerations for phenotypes****.*** The distributions of the 4 primary DDE quantitative traits (counts) were significantly right-skewed and zero-inflated (**Appendix Figure 3**). During initial model fit diagnostics, we tested multiple response transformations using linear regression models adjusted for age (in months), sex, and race/ethnicity. Based on these diagnostic criteria and preliminary genome-wide association test iterations, we determined that utilizing Pearson residuals extracted from count data models was the most statistically rigorous approach. Specifically, to account for the inherent zero-inflation, we regressed these traits on age, sex, and race/ethnicity utilizing zero-inflated negative binomial (ZINB) models. The resulting Pearson residuals were subsequently carried forward as continuous response variables in the GWA linear regression models. For analyses employing the sex-stratified framework, we generated sex-specific Pearson residuals by regressing solely on age and race/ethnicity.

*Genome-wide Association study***.** We implemented three distinct analytical approaches for genome-wide association testing (**Figure 1**):

a. **Approach 1 (main discovery)**: We carried out a discovery GWAS in the full analytical sample utilizing linear and logistic regression mixed models for the quantitative and binary traits, respectively, assuming an additive genetic model. Models were adjusted for relevant fixed effects (i.e., age, sex, race/ethnicity, and the first 8 principal components of ancestry). For quantitative traits, the ZINB-derived Pearson residuals were adjusted for the 8 principal components. SAIGE was utilized to account for sample relatedness via a genetic relationship matrix (i.e., a random effect) and to account for the imbalance between cases and non-cases where applicable (Zhou et al. 2018).
b. **Approach 2 (joint 2df test**): We investigated the genetic basis of DDE by testing the hypothesis that a genetic variant has a main and/or interaction effect on DDE using a joint 2-degree-of-freedom (2df) test. This approach leverages potential gene-sex (GxS) interactions, efficiently accounting for the heterogeneity of genetic effects between the sexes to optimize discovery power.
c. **Approach 3 (sex-stratified analysis)**: We utilized a stratified framework where the sample was split into strictly male and female strata. Genetic main-effects discovery models were performed independently within each stratum (as described in Approach 1). A conceptual schema of this modeling approach for the continuous traits (where *Y* is the Pearson residual) is as follows:

$$Female:Y= \beta_{0}^{(female)}+\beta_{G}^{(female)}\times SNP+\beta_{C}^{(female)}\times covariates+e$$

$$Male:Y= \beta_{0}^{(male)}+\beta_{G}^{(male)}\times SNP+\beta_{C}^{(male)}\times covariates+e$$

The utilization of a joint (2df) test evaluating the genetic main effect and the gene-environment (GxE) interaction terms together (*P*-joint), primarily tests the null hypothesis (H_0:_ β_G_=β_GxE_=0). This approach has been shown to yield improved statistical power, increasing the likelihood of discovering novel genetic variants (Kraft et al. 2007; Manning et al. 2011). To formally test for evidence of GxS interaction effects, we calculated *P*-values for difference (*P-diff*) between beta coefficients derived from the two sex-specific strata. The R package EasyStrata was utilized to carry out QC, generate Manhattan and quantile-quantile (Q-Q) plots, and conduct the 1df (sex-specific *P*-value), 2df (*P-joint*), and *P-diff* tests (Winkler et al. 2015).

We excluded variants with MAF<1% and R^2^<0.3, resulting in the testing of ~14.3 million autosomal SNPs (**Appendix Table 2**). Genomic inflation (λ_GC_) was assessed using Q-Q plots. To reduce the likelihood of reporting spurious associations, SNPs with an effective sample size (effN) <100 were excluded from the reporting of significant association results. A conventional genome-wide statistical significance criterion (*P* < 5×10⁻⁸) was applied to detect novel loci (Uffelmann et al. 2021), and a stringent, study-wide Bonferroni-corrected threshold (*P* < 1.25×10⁻⁹) was utilized to rigorously account for multiple testing. A SNP was considered physically independent from other signals at a linkage disequilibrium (LD) threshold of *R²* < 0.6.

**Study-wise type-I error control**

We formally interrogated 4 primary quantitative traits and 4 secondary binary traits. Additional models included gene-environment interaction terms for sex and strictly sex-stratified models evaluated for between-stratum differences. Altogether, 40 distinct GWAS models were implemented. To adequately control Type I error for novel locus discovery across the entire study, the study-wide Bonferroni correction resulted in a stringent genome-wide statistical significance threshold of *P* = 5×10⁻⁸ / 40 = 1.25×10⁻⁹.

**Functional annotation of GWAS of DDE results**

We used FUMA GWAS (Functional Mapping and Annotation of Genome-Wide Association Studies) to facilitate functional annotation of the GWAS results (Watanabe et al. 2017). Results were mapped from build hg38 to hg19 (i.e., “lift over”) prior to FUMA. The SNP2GENE process utilized tools including ANNOVAR (Wang et al. 2010), CADD scoring (Rentzsch et al. 2019), RegulomeDB scoring (Boyle et al. 2012), and 15-core chromatin state mapping (Ernst and Kellis 2012) to annotate candidate SNPs. FATHMM-XF was queried to evaluate variant pathogenicity (Rogers et al. 2018), alongside HaploReg for regulatory roles (Ward and Kellis 2012), GTEx for tissue-specific gene expression (Lonsdale et al. 2013), and GeneCards (Safran et al. 2021) to ascertain the biological function of overlapping/proximal genes. We also searched the GWAS-catalog (MacArthur et al. 2017) for previously reported associations of lead SNPs and their proxies (R^2^≥0.80).

**Gene-based test and Gene-set analysis**

MAGMA v1.6 was utilized for gene analysis within FUMA. SNPs were mapped to protein-coding genes based on Ensembl build 85, with genome-wide significance conservatively set at 0.05 / 18,273 genes tested = 2.74 x 10^-6^. Gene-level *P*-values were carried forward to gene-set analyses utilizing MSigDB v7.0 "Curated gene sets" and "GO terms".

**Pathway enrichment test**

Pathway enrichment testing evaluated the prioritized genes in the GENE2FUNC process using hypergeometric tests against pre-defined gene sets from MSigDB, WikiPathways, and the GWAS Catalog. Protein-coding genes in the FUMA database served as the background, and enrichment *P*-values were adjusted for multiple testing using the Benjamini-Hochberg False Discovery Rate (FDR) correction.

**Generalization.** Generalization of signals was evaluated using summary statistics from Alotaibi et al. (n=7,159), a multi-ethnic GWAS of enamel hypoplasia among 7–82-year-olds (Alotaibi et al. 2022) , alongside two large-scale genome-wide meta-analyses of dental caries in children (n=19,000) (Haworth et al. 2018) and adults (n=487,000) (Shungin et al. 2019). We evaluated the summary estimates of our 38 genome-wide significant SNPs for directional consistency and Bonferroni-corrected statistical significance (*P* < 1.3×10⁻³, i.e., 0.05/38). The term "generalization" (rather than "replication") is used strictly due to inherent differences in study designs, notably the lack of age overlap between cohorts, our inclusion of diverse DDE phenotypes beyond hypoplasia, and our unique modeling of gene-sex interactions. When a lead SNP was absent from an external dataset, highly correlated proxy SNPs (*R²* ≥ 0.80 in African, Admixed American, and European populations) were examined. Additionally, we reviewed regional association plots (±250Kb) from the GLIDE consortium studies (Haworth et al. 2018) (Shungin et al. 2019) and our early childhood caries (ECC) cohort (Shrestha et al. 2025) using identical index SNPs to systematically detect co-localizing signals.

**Cross-trait pleiotropy look-up in ECC**

To investigate whether the established clinical association between DDE and ECC is driven by shared genetic underpinnings, we conducted a comprehensive cross-trait pleiotropy look-up. We evaluated 939 DDE trait-variant associations against primary (cavitated lesions; ICDAS≥3) and secondary (clinically manifest; ICDAS≥1) ECC phenotypes using summary statistics from our recently reported multi-ancestry GWAS of ECC conducted within the same ZOE 2.0 cohort (Shrestha et al. 2025; n=6,103). This look-up systematically evaluated both main genetic effects and gene-environment interactions (i.e., interactions with fluoride and sugary beverage consumption frequency) to estimate the proportion of loci reaching nominal or genome-wide significance.

**APPENDIX FIGURES**

**Appendix Figure 1.** Flowchart illustrating enrollment, clinical examinations, and derivation of the analytical samples in ZOE 2.0 study, NC, USA. Adapted from Divaris et al. (Divaris et al. 2020)

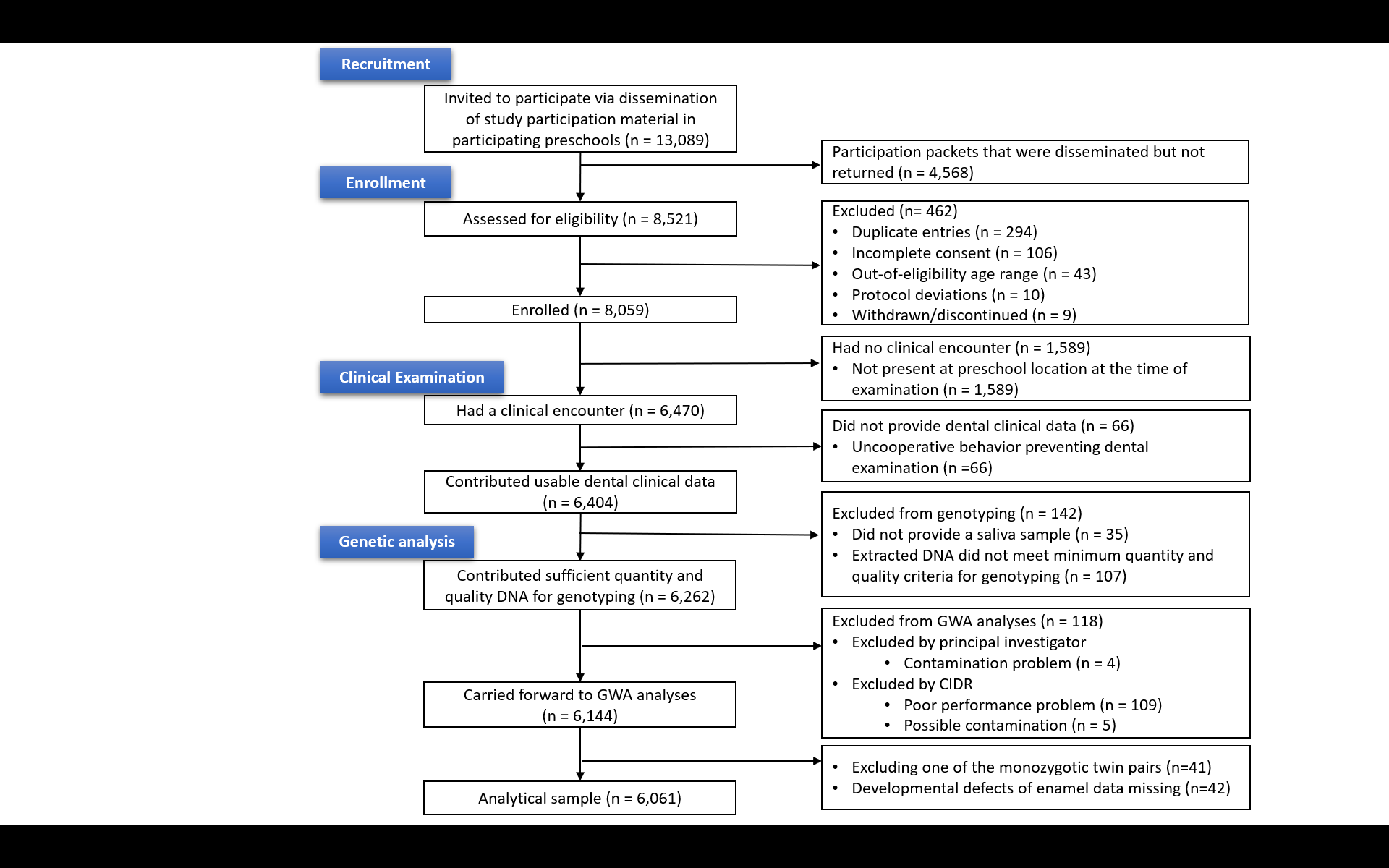

**Appendix Figure 2.** The terms, definitions and anatomy of developmental defects of enamel

| 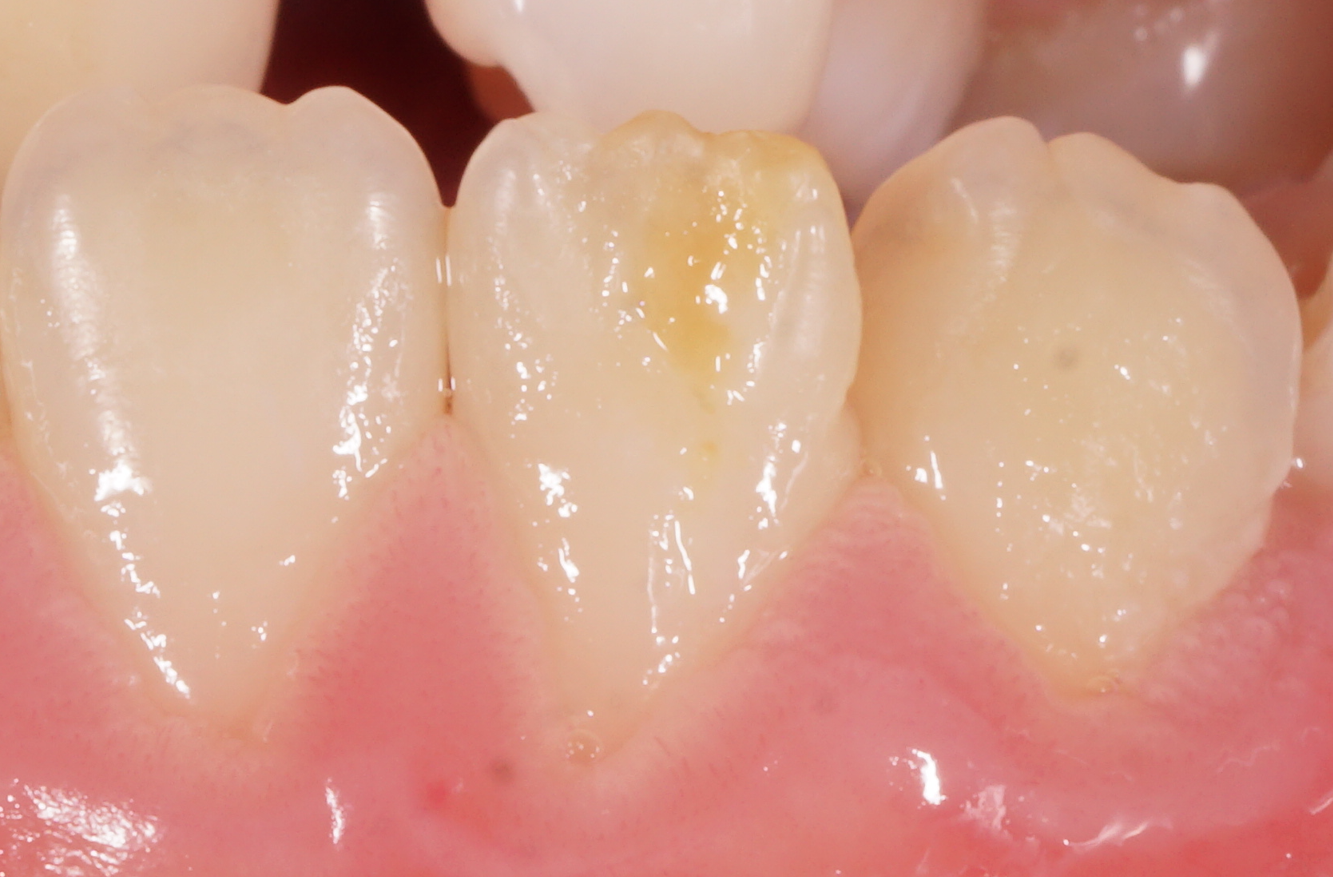 | **Hypoplasia:** Reduction in quantity of enamel tissue formed |
| --- | --- |
| 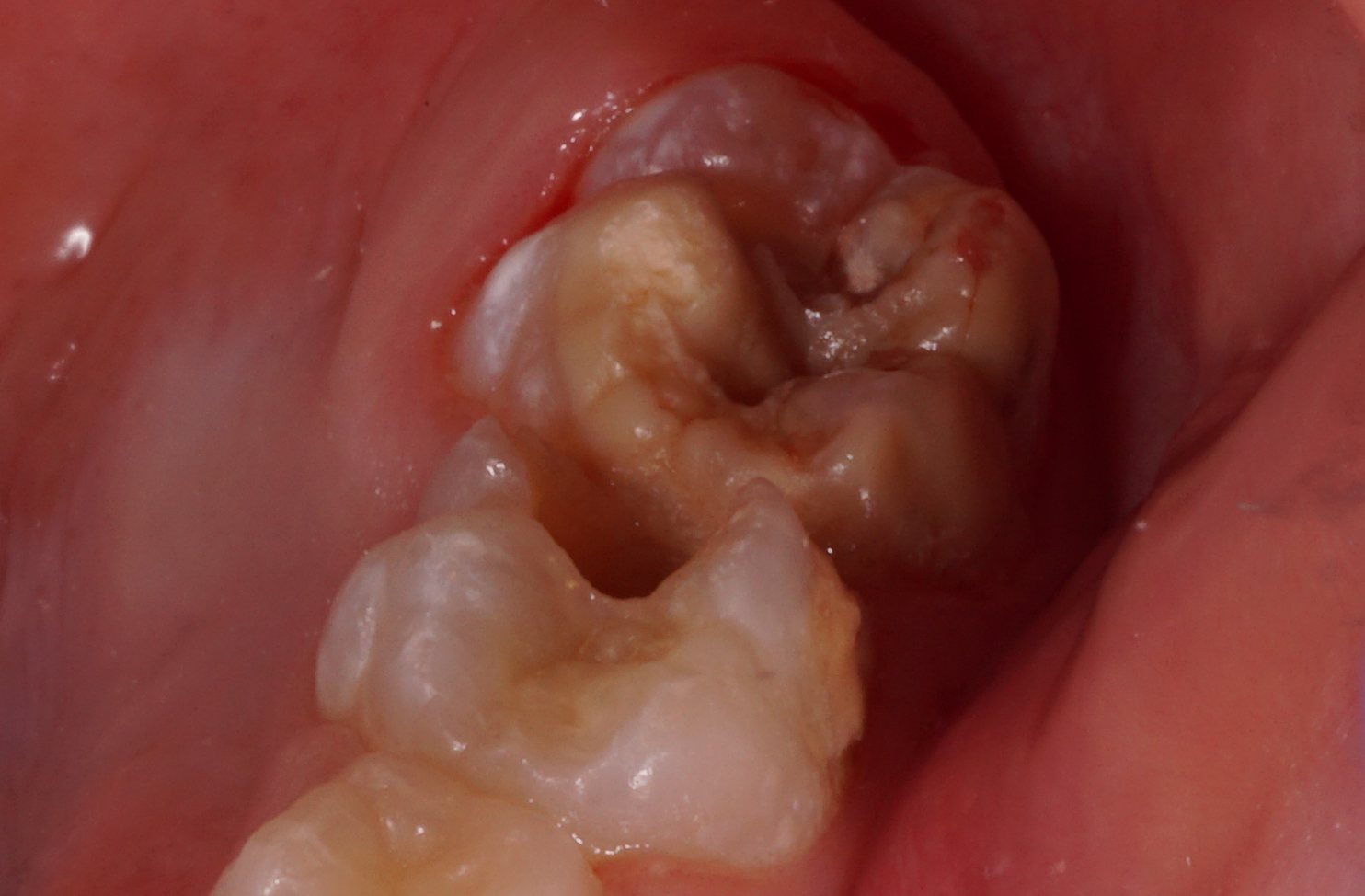 | **Demarcated opacity:** An opacity confined to a relatively small area of tooth enamel |
| 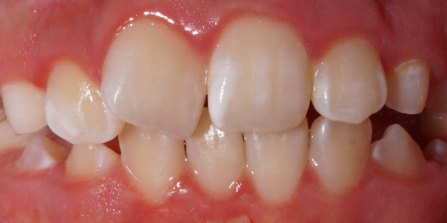 | **Diffuse opacity:** An opacity distributed over a relatively large area of tooth enamel |

**Appendix Figure 3.** Distribution of the four DDE count traits. The distributions are right skewed and zero-inflated. To account for the zero-inflation, we regressed these traits on age, sex and race/ethnicity to produce Pearson residuals using a zero-inflated negative binomial (ZINB) model. We reached a consensus that owing to the relatively large sample size and leveraging the central limit theorem, it is not necessary for these residuals to be normally distributed. Thus, we carried forward the Pearson residuals to the genome-wide association tests for this trait.

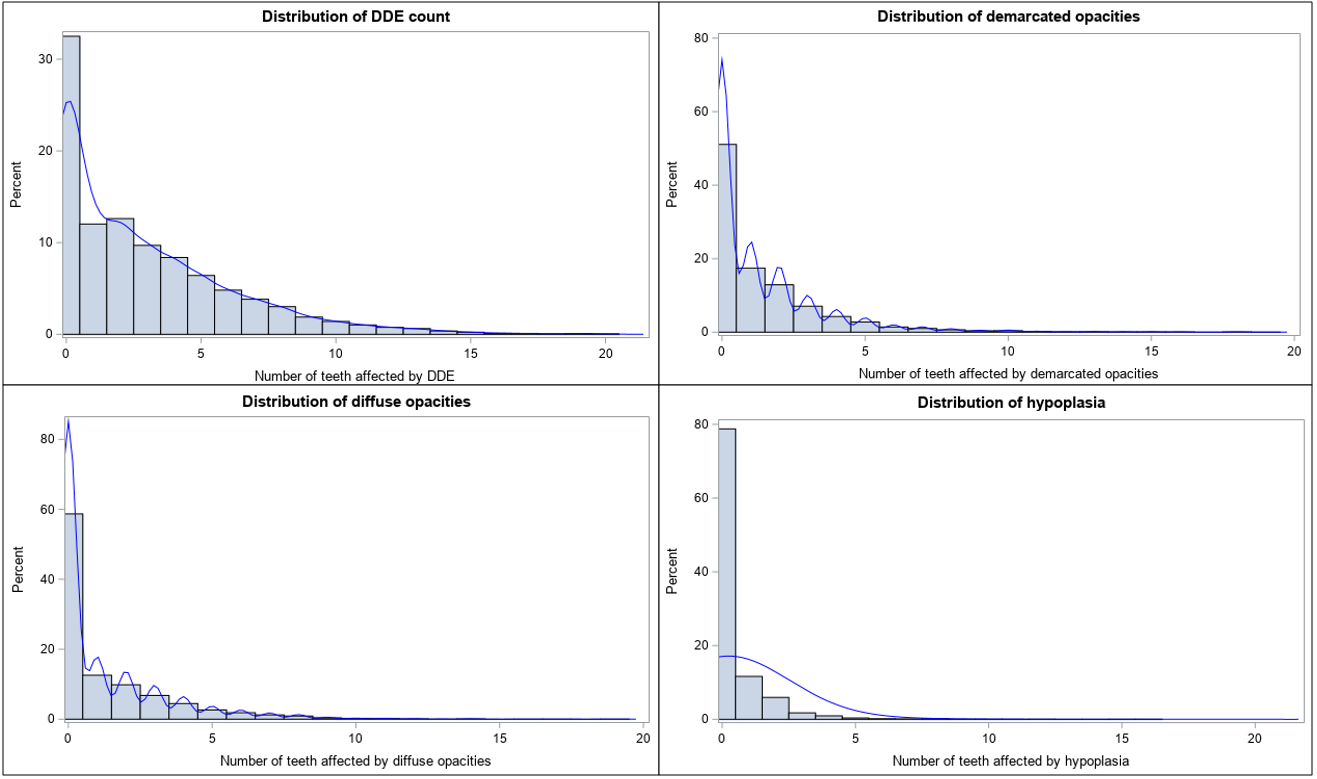

**Appendix Figure 4.** Q-Q plots for traits with genome-wide statistically significant loci in the main discovery analysis. A) DDE count, B) Demarcated opacities count, C) Diffuse Opacities count, D) Hypoplastic defects count, E) Diffuse opacities (Binary)

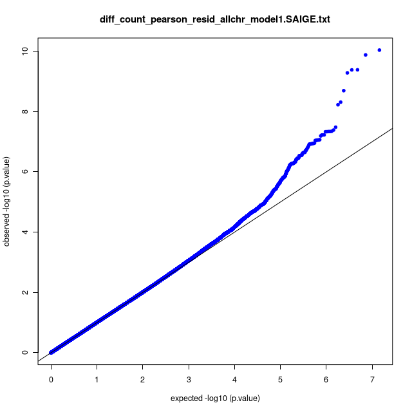

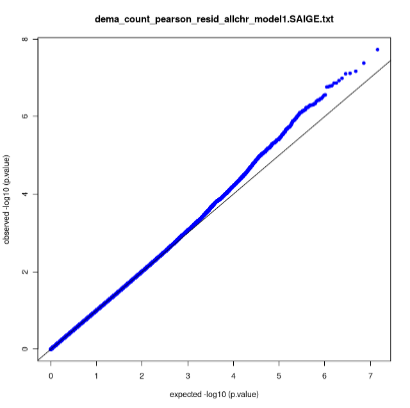

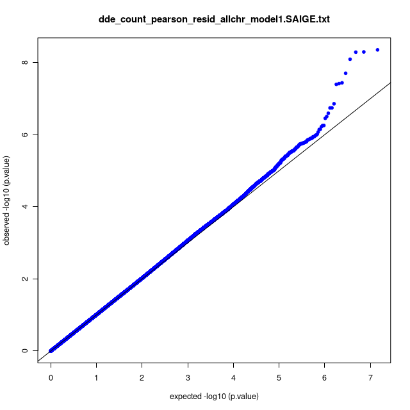

(C)

(A)

(B)

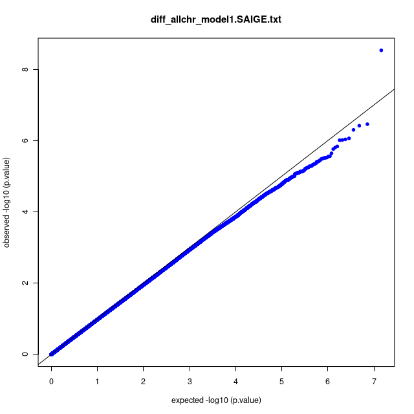

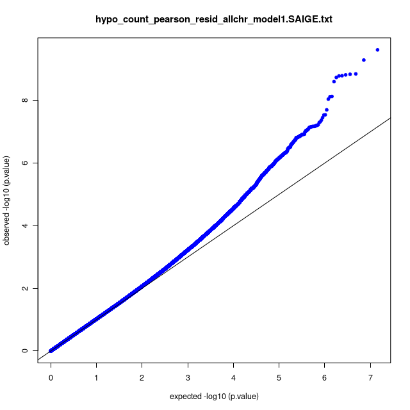

(E)

(D)

**Appendix Figure 5.** Regional association (LocusZoom) plots for the genome-wide significant loci for the combined developmental defects of enamel count trait. (A) *Y_RNA*, (B) *DEK*, (C) *RP4-724E13.2*, (D) *HBS1L*

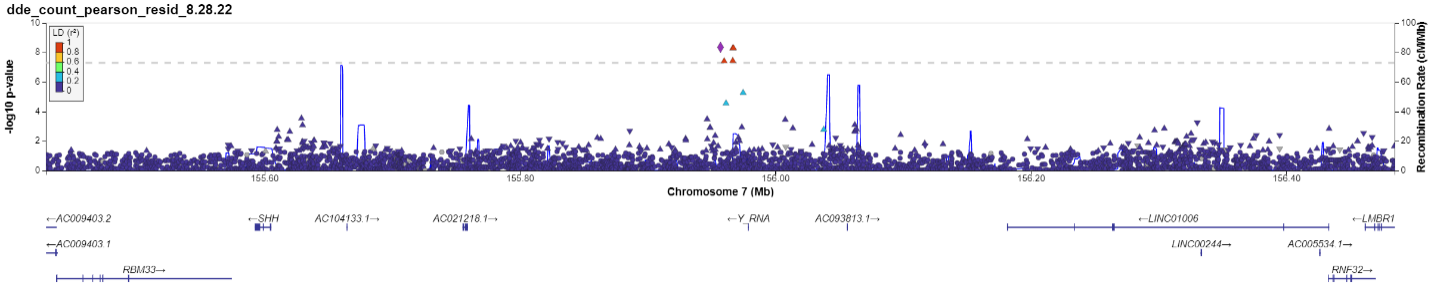

(A)

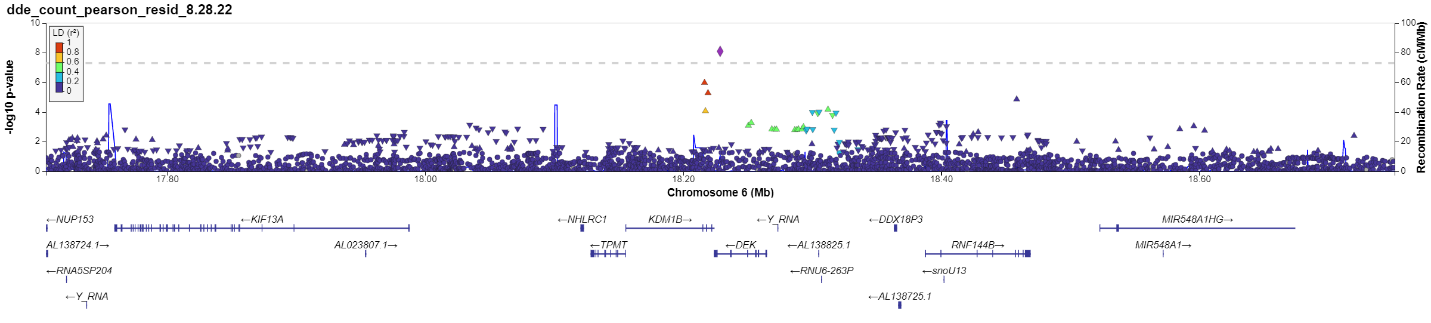

(B)

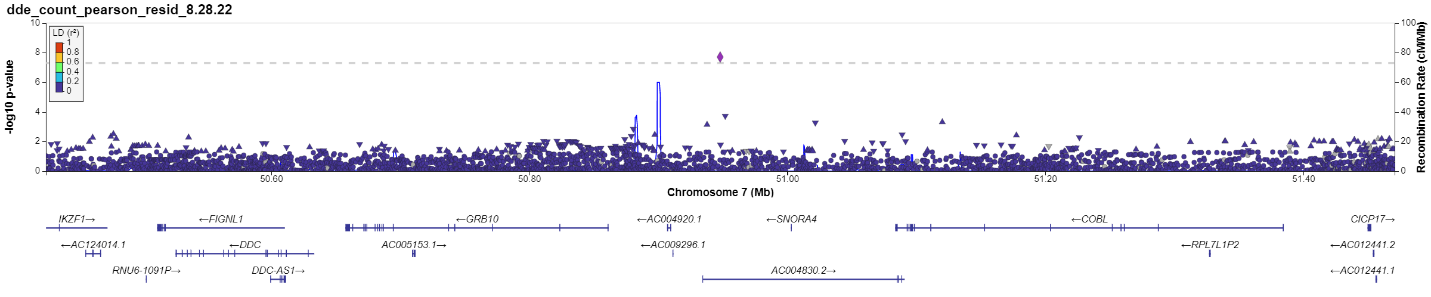

(C)

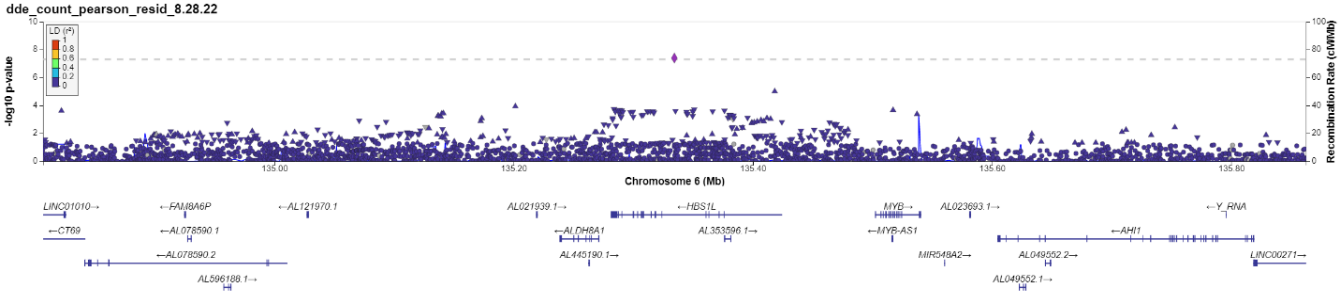

(D)

**Appendix Figure 6.** Regional association (LocusZoom) plots of two loci that emerged in the present GWAS of DDE, each within ~120 Kb of a signal reported in a related dental-trait GWAS: (1) chromosome 15 near *AC007950.1*, where the GLIDE1 adult caries (DMFS) GWAS (Shungin et al. 2019) reports a signal at p = 1.3×10⁻²⁶ (~117 Kn from the lead variant in our hypoplasia count GWAS); and (2) chromosome 6 within *DEK*, where a GWAS for clinical decay/caries (Shrestha et al. 2025) in our study population reports a signal at p = 1.8×10⁻⁷ (~113 Kb from the lead variant in our DDE count GWAS)

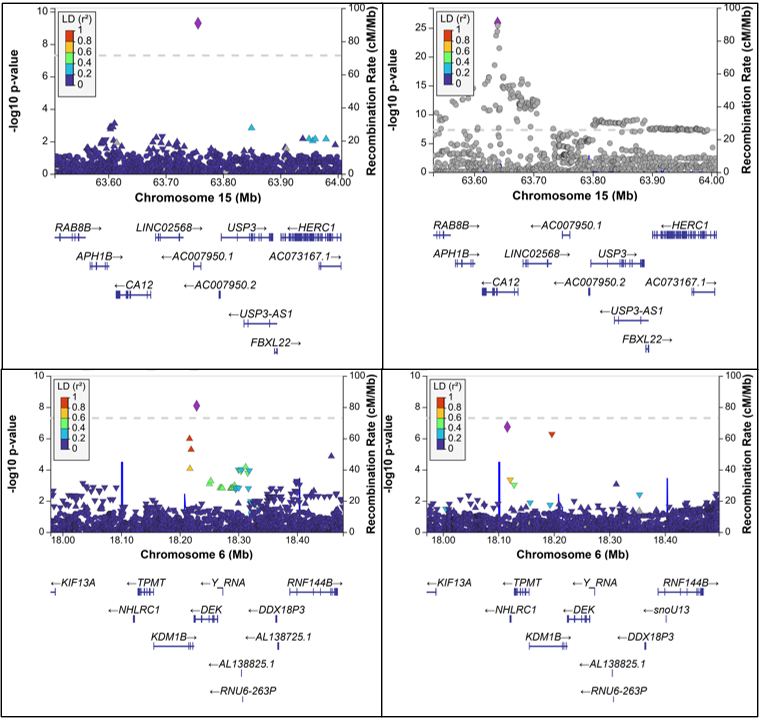

**APPENDIX TABLES**

**Appendix Table 1.** Overview of the DDE phenotypes interrogated in this study and their descriptive information in the study sample

| **Traits** | **Definition** | **Variable type** | **Mean (standard deviation) or %*** |
| --- | --- | --- | --- |
| **Primary (quantitative, count)** |  |  |  |
| Developmental defects of enamel (any) | Sum of teeth that have demarcated opacities, diffuse opacities, and hypoplastic defects. | Quantitative (Range: 0-20) | 2.9 (3.2) |
| Demarcated opacities | Sum of teeth that have demarcated opacities. | Quantitative (Range: 0-19) | 1.3 (2.1) |
| Diffuse opacities | Sum of teeth that have diffuse opacities | Quantitative (Range: 0-19) | 1.2 (2.0) |
| Hypoplastic defects | Sum of teeth that have hypoplastic defects. | Quantitative (Range: 0-16) | 0.4 (1.0) |
| **Secondary (binary, case status)** |  |  |  |
| Developmental defects of enamel (any, %) | ≥1 teeth with any form of developmental defect of enamel, in any primary tooth of a child under 6 years. | Binary | 67.5 |
| Demarcated opacities (%) | ≥1 teeth with demarcated opacities, in any primary tooth of a child under 6 years. | Binary | 48.9 |
| Diffused opacities (%) | ≥1 teeth with diffuse opacities, in any primary tooth of a child under 6 years. | Binary | 41.3 |
| Hypoplastic defects (%) | ≥1 teeth with hypoplastic defects, in any primary tooth of a child under 6 years. | Binary | 21.2 |

*Prevalence (%) for the binary traits. Mean (SD), i.e. average number of teeth affected for the quantitative traits

**Appendix Table 2.** Details of genotyping, quality control and statistical analysis

| **Genotyping** | | | | | | **Sample quality control and exclusion criteria** | | | | **Prephasing software** | **Imputation** | | | **Filters (No. of SNPs)** |
| --- | --- | --- | --- | --- | --- | --- | --- | --- | --- | --- | --- | --- | --- | --- |
| **DNA source** | **Genotyping array** | **Genotype calling algorithm** | **Inclusion criteria** | | | **Call rate per SNP** | **Call rate per participant** | **Other** | **SNPs after QC** |  | **SNPs used for imputation** | **Imputation software** | **Reference panel** | All variants (genotyped+ imputed): 307,883,017 |
|  |  |  | **MAF** | **Call rate** | **p-value (HWE)** |  |  |  |  |  |  |  |  | All non-monomorphic variants (genotyped+ imputed): 144,431,368 |
| Saliva | Infinium™ Global Diversity Array-8 v1.0 | GenomeStudio version 2011.1 | **≥**0.01 | **≥**99% | >8e-4 | >99.8% | >99% | MZ twins | 1,441,911 | Eagle2 V2.4 | 1,142,635 | Minimac4 | TOPMED Reference Panel | MAF>0.01 and R2>0.03 (Main discovery GWAS) 14,371,011 |

**Abbreviations:** MAF: Minor allele Frequency, HWE: Hardy Weinberg Equilibrium, SNP: Single nucleotide polymorphism, QC: Quality control

**Appendix Table 3.** Baseline characteristics of the study population (n=6,061), including stratification by sex.

| **Variable** | **Full sample (n=6,061)** | **Stratified sample** | |
| --- | --- | --- | --- |
|  |  | **Male (n=3,006)** | **Female (n=3,055)** |
| **Age (in months), Mean (SD)** | 53.5 (7.3) | 53.6 (7.2) | 53.4 (7.3) |
| **Gender, n (%)** |  |  |  |
| Female | 3,055 (50.4) | - | - |
| Male | 3,006 (49.6) | - | - |
| **Race/Ethnicity, n (%)*** |  |  |  |
| Non-Hispanic African American | 2,890 (47.7) | 1,387 (46.1) | 1,503 (49.2) |
| Hispanic American | 1,213 (20.0) | 634 (21.1) | 579 (19.0) |
| Non- Hispanic White | 1,068 (17.76 | 550 (18.3) | 518 (17.0) |
| American Indian/Alaskan Native | 146 (2.4) | 75 (2.5) | 71 (2.3) |
| More than one race | 631 (10.4) | 299 (10.0) | 332 (10.9) |
| Other | 109 (1.8) | 58 (1.9) | 50 (1.6) |
| **Counts / Quantitative traits, Mean (SD)*** |  |  |  |
| Developmental defects of enamel | 2.9 (3.2) | 3.0 (3.3) | 2.7 (3.1) |
| Demarcated opacities | 1.3 (2.1) | 1.4 (2.3) | 1.3 (2.0) |
| Diffuse opacities | 1.2 (2.0) | 1.3 (2.1) | 1.1 (1.9) |
| Hypoplastic defects | 0.4 (1.0) | 0.4 (1.1) | 0.4 (1.0) |
| **Binary traits, n (%)** |  |  |  |
| Developmental defects of enamel | 4,088 (67.5) | 2,073 (69.0) | 2,015 (66.0) |
| Demarcated opacities | 2,965 (48.9) | 1,476 (49.1) | 1,489 (48.7) |
| Diffuse opacities | 2,503 (41.3) | 1,312 (43.7) | 1,191 (39.0) |
| Hypoplastic defects | 1282 (21.2) | 672 (22.4) | 610 (20.0) |
| **Abbreviations**. SD: Standard deviation; n: number; %: Percentage  * Missing data: Race/Ethnicity: 4; DDEs: 42 | |  |  |

**Appendix Table 4.** SNP-based heritability (*h*^2^) estimated using imputed data among unrelated individuals (n=5,580) and Concordance estimates (95% confidence interval) among pair of related individuals at different levels of relatedness (assessed using the kinship coefficient) for the DDE traits

|  | **Heritability estimates** | | **Concordance Estimates** | | |
| --- | --- | --- | --- | --- | --- |
| **DDE traits** | ***h*^2^* (SE)** | ***P*-value** | **Monozygotic twins (41 pairs)** | **1^st^ degree relatives (259 pairs)** | **2^nd^ & 3^rd^ degree relatives (382 pairs)** |
| **Counts/Quantitative (number of affected teeth)** | | | | | |
| Developmental defects of enamel | 0.20 (0.06) | 7.9x10^-4^ | 0.43 [0.15, 0.65] | 0.41 [0.30, 0.50] | 0.19 [0.09, 0.28] |
| Demarcated opacities | 0.09 (0.06) | 7.3x10^-2^ | 0.03 [-0.28, 0.33] | 0.28 [0.17, 0.39] | 0.13 [0.03, 0.22] |
| Diffuse opacities | 0.16 (0.07) | 7.4x10^-3^ | 0.76 [0.59, 0.86] | 0.35 [0.23, 0.45] | 0.10 [-0.00, 0.20] |
| Hypoplastic defects | 0.07 (0.06) | 1.2x10^-1^ | 0.03 [-0.28, 0.33] | 0.02 [-0.11, 0.14] | -0.01 [-0.11, 0.09] |
| **Binary case status (any versus no affected tooth)** | | | | | |
| Developmental defects of enamel | 0.05 (0.06) | 2.4x10^-1^ | 0.03 [-0.28, 0.33] | 0.39 [0.28, 0.51] | 0.18 [0.07, 0.28] |
| Demarcated opacities | 0.07 (0.06) | 1.3x10^-1^ | 0.06 [-0.24, 0.37] | 0.29 [0.18, 0.41] | 0.12 [0.02, 0.22] |
| Diffuse opacities | 0.04 (0.06) | 3.0x10^-1^ | 0.70 [0.48, 0.92] | 0.38 [0.27, 0.49] | 0.19 [0.09, 0.29] |
| Hypoplastic defects | 0.06 (0.06) | 1.8x10^-1^ | 0.07 [-0.25, 0.39] | 0.06 [-0.06, 0.19] | 0.01 [-0.09, 0.11] |

*Adjusted for age, sex, race (6-category) and 8 ancestry principal components. All count/quantitative traits are Pearson residualized.

Note: For categorical traits, Cohen's *kappa* is calculated. For quantitative traits, intraclass correlation coefficients (ICC) are calculated using a one-way random effects model. 95% confidence intervals are presented in brackets.

**Appendix Table 5.** Genomic inflation factor (λ_GC_) for genome-wide analysis performed in this study

| **DDE traits** | **SNPs** | **λ_GC_** | **SNPs** | **λ_GC_** |
| --- | --- | --- | --- | --- |
| **Main discovery analysis** | | | | |
|  | **Quantitative** | | **Binary case status** | |
| Developmental defects of enamel | 14363837 | 1.00 | 14363837 | 0.97 |
| Demarcated opacity | 14363837 | 1.00 | 14363837 | 0.98 |
| Diffuse opacity | 14363837 | 1.01 | 14363837 | 0.97 |
| Hypoplastic defects | 14363837 | 1.03 | 14363837 | 1.00 |
| **Sex-stratified analysis** | | | | |
| **Developmental defects of enamel** | **Quantitative** | | **Binary case status** | |
| GWAS - Male | 13791004 | 1.00 | 9470591 | 1.01 |
| GWAS - Female | 13941451 | 1.01 | 9948308 | 1.01 |
| GWAS - p-joint | 13518925 | 1.00 | 9364395 | 1.00 |
| GWAS - p-diff | 13518925 | 0.99 | 9364395 | 1.00 |
| **Demarcated opacity** |  |  |  |  |
| GWAS - Male | 13791004 | 1.04 | 11529100 | 1.00 |
| GWAS - Female | 13941451 | 1.00 | 11313687 | 1.02 |
| GWAS - p-joint | 13518925 | 1.02 | 11078625 | 1.00 |
| GWAS - p-diff | 13518925 | 1.01 | 11078625 | 1.00 |
| **Diffuse opacity** |  |  |  |  |
| GWAS - Male | 13791004 | 0.99 | 10779415 | 0.99 |
| GWAS - Female | 13941451 | 1.01 | 10486195 | 1.01 |
| GWAS - p-joint | 13518925 | 1.00 | 10302996 | 0.99 |
| GWAS - p-diff | 13518925 | 0.99 | 10302996 | 1.00 |
| **Hypoplastic defects** |  |  |  |  |
| GWAS - Male | 13791004 | 1.00 | 8376258 | 1.00 |
| GWAS - Female | 13941451 | 0.93 | 8236834 | 1.00 |
| GWAS - p-joint | 13518925 | 0.93 | 8017634 | 0.98 |
| GWAS - p-diff | 13518925 | 0.95 | 8017634 | 0.99 |

**Abbreviations:** DDE: Developmental defects of enamel, SNP: Single Nucleotide Polymorphism, λ_GC_: Genomic inflation factor, GWAS: Genome-wide association study, P*_joint_*: Joint 2-degree-of-freedom test, P*_diff_*: test of difference between stratum-specific beta-coefficients

**Appendix Table 6.** Annotation of novel loci from the GWAS for developmental defects of enamel identified in Approach 1

| **Locus** | **Lead SNP (rsid)** | **Functional  annotion** | **CADD score^a^** | **RDB Score^b^** | **FATHMM-XF score^c^** | **Promoter Histone markers^d^** | **Enhancer histone markers^d^** | **DNase (Tissue or cell lines)^d^** | **Protein bound^d^** | **Motifs changes^d^** |
| --- | --- | --- | --- | --- | --- | --- | --- | --- | --- | --- |
| **Developmental defects of enamel (quantitative)** | | | | | | | | | | |
| *Y_RNA* | rs143210152 | intergenic | 0.32 | 7 | 0.062584, benign |  |  |  |  | GATA,HEY1 |
| -- | rs567243779^e^ | intronic | 0.12 | NA | 0.017215, benign (high conf.) |  |  |  |  |  |
| *RP4-724E13.2* | rs150075871 | ncRNA_intronic | 0.88 | 5 | 0.027902, benign (high conf.) |  |  |  |  | NRSF |
| *HBS1L* | rs116989668 | intronic | 2.32 | 5 | 0.04135, benign |  | LNG, BLD, STRM, FAT, MUS, SKIN, HRT, BONE, THY, BRN |  |  |  |
| **Demarcated opacities (quantitative)** | | | | | | | | | | |
| *RP11-93B14.5* | rs75880235 | exonic | 10.82 | 4 | 0.006922, benign (high conf.) | SKIN, BRN, GI, LIV | SKIN, BRST, BRN, FAT, MUS, HRT, GI, ADRL, PLCNT, PANC, SPLN, BLD, LNG, ESDR | SKIN, GI, LNG, PANC, LIV, BRST | ERALPHA_A | ERalpha-a,Zbtb3 |
| -- | rs895438538^e^ | intergenic | 0.99 | NA | NA |  |  |  |  |  |
| **Diffuse opacities (quantitative)** | | | | | | | | | | |
| *Y_RNA* | rs77669438 | intergenic | 0.12 | 6 | 0.047708, benign |  |  |  |  | 4 altered motifs |
| -- | rs1658697761^e^ | intergenic | 1.14 | NA | NA |  |  |  |  |  |
| **Hypoplastic defects (quantitative)** | | | | | | | | | | |
| *RP11-157E21.1* | rs192067416 | intergenic | 1.99 | 6 | 0.071444, benign |  |  |  |  | AP-2, BCL, EBF, Egr-1, Ets, Hic1, Myc, Rad21, Znf143 |
| *AC007950.1* | rs79505735 | ncRNA_intronic | 10.51 | 7 | 0.029107, benign (high conf.) |  | ESDR, BRN, MUS, VAS, GI, PANC, LNG | GI |  | Bcl6b |
| *RP1-167G20.1* | rs149507176 | intergenic | 6.39 | 6 | 0.066973, benign |  |  |  |  | BATF, Irf, Nkx2, Nkx3, STAT |
| *ADAMTSL1* | rs115373518 | intergenic | 1.74 | 7 | 0.061862, benign |  | THYM |  |  | PLAG1 |
|  | -- | intronic | 1.65 | NA | NA |  |  |  |  |  |
| *FHIT* | rs73835238 | intronic | 0.52 | 5 | 0.037821, benign |  |  |  | KAP1 | Nanog, Pou2f2, Pou5f1, RXRA, TATA, YY1 |
| *UBE2G2* | rs115617269 | intergenic | 0.03 | 5 | 0.059699, benign |  | ESC, IPSC, ESDR, BLD, SKIN, GI, BRST |  |  | Ets, GR, HEY1, NF-kappaB, NRSF, TATA, YY1, p300 |
| *MPV17L2* | rs79028494 | intergenic | 2.73 | 5 | 0.06986, benign |  | ESC, IPSC, LIV, BLD | ESC, IPSC, KID, BLD |  |  |
| *PPAPDC3* | rs75061854 | intronic | 0.07 | 5 | 0.022962, benign (high conf.) | HRT, MUS | IPSC, MUS, HRT, ADRL | HRT |  | CAC-binding-protein,CTCF |
| *CLUL1* | rs114989924 | intronic | 13.03 | 2b | 0.197737, benign |  | ESDR |  |  | Nanog,Pou2f2,TATA |
| *MAML2* | rs150489138 | intronic | 0.35 | 6 | 0.02615, benign (high conf.) |  | BLD |  |  | Foxm1, GATA, SIX5, Smad, Sox, Znf143 |
| *MTHFD1L* | rs11963290 | intronic | 0.10 | 6 | 0.024124, benign (high conf.) |  |  |  |  | CACD, Klf4, Klf7, NRSF, SP1 |
| **Diffuse opacities (binary)** | | | | | | | | | | |
| *PPIL4* | rs184135739 | intergenic | 0.44 | 7 | 0.059911, benign | BLD | BLD, MUS |  |  | BCL, Eralpha-a, Irf, SP1, ZBRK1 |

**Abbreviations**: Chr - chromosome; EA - effect allele; OA - Other allele; SNP - Single nucleotide polymorphism; RDB – Regulome DB

a) Scores the deleteriousness of the single nucleotide variants and INDELs in human genome (PMID: 30371827).
b) http://regulomedb.org; Likely to affect binding and linked to expression of a gene target (1a:eQTL + TF binding + matched TF motif + matched DNase Footprint + DNase peak, 1b:eQTL + TF binding + any motif + DNase Footprint + DNase peak, 1c:eQTL + TF binding + matched TF motif + DNase peak, 1d: eQTL + TF binding + any motif + DNase peak, 1e:eQTL + TF binding + matched TF motif; 1f:eQTL + TF binding / DNase peak); Likely to affect binding (2a: TF binding + matched TF motif + matched DNase Footprint + DNase peak, 2b:TF binding + any motif + DNase Footprint + DNase peak, 2c:TF binding + matched TF motif + DNase peak); [PMID: 22955989]
c) Based on Enhanced Accuracy in Predicting the Functional Consequences of Non-Coding and Coding Single Nucleotide Variants (FATHMM-XF) score PMID: 28968714
d) Promoter and Enhancer histone markers, Dnase, protein bound, and motif changes based on HaploReg v4.1 information (http://www.broadinstitute.org/mammals/haploreg/haploreg.php) that is derived from Roadmap Epigenomic Project and ENOCDE project .
e) Not found in Haploreg V4.1

**Appendix Table 7.** Annotation of novel loci from the GWAS for developmental defects of enamel identified in Approach 2 and 3

| **Locus** | **Functional  annotation** | **rsid** | **CADD score^a^** | **RDB Score^b^** | **FATHMM-XF score^c^** | **Promoter Histone markers^d^** | **Enhancer histone markers^d^** | **DNase (Tissue or cell lines)^d^** | **Protein bound^d^** | **Motifs changes^d^** |
| --- | --- | --- | --- | --- | --- | --- | --- | --- | --- | --- |
| **Quantitative traits** | | | | | | | | | | |
| **Developmental defects of enamel - Female stratum** | | | | | | | | | | |
| *ALDH1A1*** | none | rs141753205 | 2.34 | 7 | 0.045185, benign | LNG | SKIN, GI, PANC, LIV | HRT, GI |  | NRSF |
| *RP11-384F7.1* | none | rs202021524 | 1.20 | NA | Error |  |  |  |  | Elf3, Myc, PAX-5, Pou2f2, Tel2, p300 |
| **Developmental defects of enamel - Male stratum** | | | | | | | | | | |
| *RP11-415K20.1#* | none | rs16826979 | 0.70 | 7 | 0.114455, benign |  | ESDR, FAT, BLD | ESDR |  | Bbx, DMRT7, HP1, Hbp1 |
| *TMPRSS13* | none | rs117786109 | 0.43 | 5 | 0.097285, benign |  | BLD, THYM | BLD |  |  |
| **Developmental defects of enamel - Joint 2df test** | | | | | | | | | | |
| *Y_RNA* | none | rs143210152 | 0.32 | 7 | 0.062584, benign |  |  |  |  | GATA, HEY1 |
| **Demarcated opacities - Female stratum** | | | | | | | | | | |
| *JRKL:JRKL-AS1* | none | rs141989840 | 6.95 | 7 | 0.0319, benign |  |  |  |  | HDAC2, Sin3Ak-20 |
| *RP11-534P19.1**#* | none | rs61471572 | 1.79 | 7 | 0.068914, benign |  |  |  |  |  |
| *DLGAP2* | intronic | rs2906574 | 2.13 | 5 | 0.014629, benign (high conf.) |  |  |  |  | CTCF, TCF12 |
| *FHIT#* | intronic | rs72891211 | 2.20 | 6 | 0.042629, benign |  |  |  |  | Hsf, Pax-4, Pou2f2, Pou3f2, RXRA, p300 |
| **Demarcated opacities - Male stratum** | | | | | | | | | | |
| *PTPRK**#* | intronic | rs77758675 | 10.62 | 4 | 0.00743, benign (high conf.) | BLD, BRN, FAT, MUS |  | 31 tissues | POL2, TAF1 | ATF3, CTCF, HEY1, SRF, Znf143 |
| *RERE**#* | intronic | rs77921067 | 0.49 | 6 | 0.043843, benign | ESDR | 19 tissues |  |  | Barx2, CDP, En-1, Msx-1, Sox |
| *ZNRF3**#* |  | rs369163187 | 3.54 | 7 | 0.022277, benign (high conf.) | Not found in 1000G Phase I |  |  |  |  |
| *RP11-57H14.3*** | none | rs77466522 | 9.94 | 4 | 0.103966, benign |  | 12 tissues | LNG, CRVX, BLD | GR |  |
| *EFR3B**#* | intronic | rs188016617 | 3.00 | 7 | 0.02013, benign (high conf.) |  |  |  |  | STAT |
| *FRMD5#* | intronic | rs146484655 | 1.29 | 6 | 0.027495, benign (high conf.) |  |  |  |  | AP-1, Zfx |
| *RP5-963E22.4#* | none | rs201297757 | 0.49 | NA | Error |  |  |  |  | AP-2, BDP1.BHLHE40, CTCF, EBF, Myc, NRSF, Rad21, YY1, Zic |
| *LINC01508#* |  | rs569532598 | 7.95 | 7 | 0.040741, benign | Not found in 1000G Phase I |  |  |  |  |
| **Demarcated opacities - p-diff** | | | | | | | | | | |
| *SLC2A3* | none | rs61921622 | 1.69 | 2b | 0.07608, benign |  | SKIN | 13 tissues |  | HNF4, Pax-5, Zbtb3 |
| **Diffuse opacities - Female stratum** | | | | | | | | | | |
| *GBE1*** | intronic | rs57583981 | 0.07 | 6 | 0.027666, benign (high conf.) |  |  |  |  | Arid3a, Dbx1, HNF1, Hoxd10, Ncx |
| *TENM2**^ⴕ^* | intronic | rs141872488 | 0.82 | 5 | 0.025738, benign (high conf.) |  | LIV | IPSC |  |  |
| *THBS4#* | none | rs77790627 | 1.26 | 5 | 0.027626, benign (high conf.) |  | 11 tissues |  |  | SIX5 |
| *RP11-384F7.1* |  | -- | 1.20 | NA | Error | Not found |  |  |  |  |
| **Diffuse opacities - Male stratum** | | | | | | | | | | |
| *RP11-473I1.6#* | none | rs140095098 | 0.15 | 6 | 0.051078, benign |  | PLCNT |  |  |  |
| **Diffuse opacities - Joint 2df test** | | | | | | | | | | |
| *Y_RNA* | none | rs77669438 | 0.12 | 6 | 0.047708, benign |  |  |  |  | AP-1, BHLHE40, ELF1, Myc |
| *CTA-21C21.1* | none | rs11440629 | 1.14 | NA | Error |  |  |  |  | AP-3, Bbx, CEBPG, HNF6, Hoxa5, Pbx-1 |
| **Hypoplastic defects - Female stratum** | | | | | | | | | | |
| *ADAMTSL1**#* | none | rs55947174 | 6.21 | 5 | 0.252397, benign |  | ESC, ESDR, IPSC, THYM | ESC, IPSC |  | Evi-1, HDAC2 |
| *KRTAP7-1*** | none | rs111896551 | 3.65 | 5 | 0.122033, benign |  |  |  |  | CEBPB, Eomes, PLZF, TCF4, p300 |
| *ADD2**#* |  | rs146459483 | 13.62 | 7 | 0.045093, benign |  |  |  |  |  |
| *CNTN4#* |  | -- |  |  | Error | Not found |  |  |  |  |
| *STARP1**#* |  | rs973279911 | 1.91 | NA | Error | Not found in 1000G Phase I |  |  |  |  |
| *HPD**#* |  | rs568971993 | 8.51 | NA | 0.060086, benign | Not found in 1000G Phase I |  |  |  |  |
| *TMEM40**#* | none | rs180978195 | 6.73 | 6 | 0.021703, benign (high conf.) |  | ESDR, SKIN, BRST, MUS, PLCNT |  |  | Mef2, SIX5 |
| *GABRG3**#* | none | rs191103614 | 1.69 | 7 | 0.019089, benign (high conf.) |  |  |  |  |  |
| *CCSER1*** | none | rs139393106 | 3.94 | 7 | 0.076176, benign |  |  |  |  | PRDM1, Pax-4, RXRA, TBX5, p300 |
| *ZNF331#* | intronic | rs192307826 | 13.65 | 7 | 0.038622, benign |  |  |  |  | Ets, Znf143, p300 |
| *JRKL:JRKL-AS1**#* | none | rs11826357 | 6.25 | 5 | 0.022587, benign (high conf.) | MUS | ESC, IPSC, ESDR, BRN, FAT, HRT, KID | BRN, MUS, HRT, CRVX |  | CEBPA, Nanog, STAT, p300 |
| *MMP26* | none | rs17251629 | 0.37 | 7 | 0.028607, benign (high conf.) |  | BLD |  |  |  |
| *CTD-2021A8.3**#* | none | rs140671047 | 0.59 | 6 | 0.058629, benign |  |  |  |  | BRCA1, FAC1, Foxp1, Pou5f1 |
| *MCTP2#* | none | rs117304168 | 0.81 | 5 | 0.027902, benign (high conf.) | BLD | BLD, FAT, BRST, GI, SKIN | BLD, BRST, SKIN |  | Foxa, NF-I, p300 |
| *ZNF823#* |  | -- | 1.63 | NA | Error | Not found |  |  |  |  |
| *CYP4F22#* | none | rs149762022 | 2.66 | 7 | 0.070557, benign |  | LIV |  |  |  |
| *PLEKHD1* | intronic | rs147589787 | 1.66 | 6 | 0.027401, benign (high conf.) |  |  |  |  | Ik-3, STAT |
| *MAST4* |  | rs1753671813 | 0.39 | NA | Error | Not found in 1000G Phase I |  |  |  |  |
| *NF1* | intronic | rs147836184 | 2.29 | 7 | 0.026501, benign (high conf.) | BRN | IMR90, ESC, IPSC, ESDR, BRST, BRN, MUS, VAS | ESDR |  | AP-1, Hic1, Znf143 |
| **Hypoplastic defects - Male stratum** | | | | | | | | | | |
| *AC073928.2**#* |  | rs141261078 | 0.03 | 5 | 0.094377, benign |  |  |  |  | PU.1 |
| *GADL1**#* | none | rs144577905 | 0.24 | 5 | 0.100679, benign |  |  |  |  |  |
| *PDCD1LG2**^ⴕ#^* | intronic | rs138482896 | 3.06 | 7 | 0.040052, benign | BLD | BLD, GI, SPLN | BLD |  | AP-1, Evi-1, Myf, ZEB1 |
| *RP11-586E1.1**#* | none | rs115583696 | 3.11 | 6 | 0.135223, benign |  |  |  |  | ELF1, Gfi1, NF-kappaB, Nanog, PU.1 |
| *RAVER2**#* |  | -- | 1.65 | NA | Error | Not found |  |  |  |  |
| *PIK3R4#* | 5'-UTR | rs150504962 | 10.95 | NA | Error | 24 tissues |  | 15 tissues |  | DMRT4, Eomes, LXR |
| *CCDC18* | intronic | rs187188755 | 1.46 | 6 | 0.029739, benign (high conf.) |  |  |  |  | Dbx1, Mef2, Pax-4, TATA, Zfp105 |
| *BCHE#* | intronic | rs6805552 | 2.85 | 7 | 0.013525, benign (high conf.) |  |  |  |  | Pax-4 |
| *AGTRAP#* | intronic | rs118165546 | 0.85 | 4 | 0.025802, benign (high conf.) |  | 10 tissues | 4 tissues |  | AP-2rep, CTCF, SP1 |
| *KRTAP9-6#* |  | rs2016470489 | 0.09 | 7 | 0.280171, benign | Not found in 1000G Phase I |  |  |  |  |
| *WI2-81516E3.1#* | none | rs146461752 | 0.70 | 6 | 0.058654, benign |  |  | IPSC, BLD |  | Maf, SIX5 |
| *TENM2* | intronic | rs62383285 | 2.05 | 5 | 0.018536, benign (high conf.) |  | IPSC | BRN, SKIN, LNG |  |  |
| **Hypoplastic defects - Joint 2df test** | | | | | | | | | | |
| *AC007950.1* | none | rs79505735 | 10.51 | 7 | 0.029107, benign (high conf.) |  | ESDR, BRN, MUS, VAS, GI, PANC, LNG | GI |  | Bcl6b |
| *RP1-167G20.1* | none | rs149507176 | 6.39 | 6 | 0.066973, benign |  |  |  |  | BATF, Irf, Nkx2, Nkx3, STAT |
| *RP11-157E21.2* | none | rs192067416 | 1.99 | 6 | 0.071444, benign |  |  |  |  | AP-2, BCL, EBF, Egr-1, Ets, Hic1, Myc, Rad21, Znf143 |
| *CCDC82#* | intronic | rs192993105 | 6.25 | 5 | 0.022943, benign (high conf.) |  |  |  |  |  |
| **Binary traits** | | | | | | | | | | |
| **Developmental defects of enamel - Male stratum** | | | | | | | | | | |
| *SLC19A1* | none | rs62214342 | 2.01 | 5 | 0.053637, benign | SKIN | BRN, MUS | IPSC |  | 15 altered motifs |
| **Developmental defects of enamel - p-diff** | | | | | | | | | | |
| *snoU13* | none | rs7228955 | 0.16 | 6 | 0.065299, benign |  | BLD |  |  | Crx, Pitx2 |
| **Demarcated opacities - Male stratum** | | | | | | | | | | |
| *CTD-2644I21.1*** | none | rs2776544 | 14.43 | NA | 0.112114, benign | 13 tissues | ESDR, BLD, BRN, PANC, SKIN, MUS, GI, PLCNT, ADRL | IPSC |  | EWSR1-FLI1, Pou1f1, Pou3f2 |
| *RP11-203H2.1* | none | rs17143504 | 0.88 | 7 | 0.066248, benign |  |  |  |  | Bbx, FXR, Hbp1 |
| **Diffuse opacities - Male stratum** | | | | | | | | | | |
| *LINC00670* | intronic | rs11078084 | 4.13 | 7 | 0.030831, benign |  |  |  |  | BDP1, Egr-1, Zbtb3 |

** SNP is also genome-wide statistically significant for the joint test (main genetic effect + gene x environment effect)

ⴕ SNP is also genome-wide statistically significant for the test of difference (i.e. interaction effect)

#Excluded from the results owing to the effective N (effN)<100.

**Abbreviations:** Chr - chromosome; EA: Effect allele; OA: Other allele; EAF: effect allele frequency; SNP: Single nucleotide polymorphisms; RDB: Regulome DB; 2df: 2-degree-of-freedom, P*_diff_*: p-value for difference between betas

a) Scores the deleteriousness of the single nucleotide variants and INDELs in human genome (PMID: 30371827).

b) http://regulomedb.org; Likely to affect binding and linked to expression of a gene target (1a:eQTL + TF binding + matched TF motif + matched DNase Footprint + DNase peak, 1b:eQTL + TF binding + any motif + DNase Footprint + DNase peak, 1c:eQTL + TF binding + matched TF motif + DNase peak, 1d: eQTL + TF binding + any motif + DNase peak, 1e:eQTL + TF binding + matched TF motif; 1f:eQTL + TF binding / DNase peak); Likely to affect binding (2a: TF binding + matched TF motif + matched DNase Footprint + DNase peak, 2b:TF binding + any motif + DNase Footprint + DNase peak, 2c:TF binding + matched TF motif + DNase peak); [PMID: 22955989]

c) Based on Enhanced Accuracy in Predicting the Functional Consequences of Non-Coding and Coding Single Nucleotide Variants (FATHMM-XF) score PMID: 28968714

d) Promoter and Enhancer histone markers, Dnase, protein bound, and motif changes based on HaploReg v4.1 information (http://www.broadinstitute.org/mammals/haploreg/haploreg.php) that is derived from Roadmap Epigenomic Project and ENOCDE project

e) Variant did not upload to FUMA, not found in 1000 Genomes data, with no information in HaploReg

**Appendix Table 8.** Genome-wide significant genes from gene analysis using MAGMA (v1.6)

| **Primary Trait** | **GENE** | **CHR** | **START** | **STOP** | **NSNPS** | **NPARAM** | **N** | **ZSTAT** | **P** | **SYMBOL** |
| --- | --- | --- | --- | --- | --- | --- | --- | --- | --- | --- |
| **Diffuse opacities (quantitative) - p-diff** | ENSG00000040731 | 5 | 24487209 | 24645087 | 1034 | 85 | 6057 | 4.61 | 2.0x10^-6^ | *CDH10* |

*Input SNPs were mapped to 18,273 protein coding genes. Genome wide significance was defined at *P* = 0.05/18273 = 2.74x10^-6^.

**Abbreviations**: P*_diff_*: p-value for the difference of the stratum-specific beta coefficients

**Appendix Table 9.** Significant gene sets with P_bon_<0.05 using the full distribution of SNP p-values

| **Gene Set** | **N genes** | **Beta** | **Beta STD** | **SE** | **P** | **P_bon_** |
| --- | --- | --- | --- | --- | --- | --- |
| **Trait / Analysis: Hypoplastic defects (Quantitative) / Main analysis** | | | | | | |
| Curated_gene_sets:valk_aml_with_t_8_21_translocation | 4 | 1.9 | 0.03 | 0.39 | 7.3E-07 | 1.1E-02 |
| **Trait / Analysis: Developmental defects of enamel (Quantitative) / p-difference (Sex Stratified)** | | | | | | |
| Curated_gene_sets:pid_fanconi_pathway | 44 | 0.6 | 0.03 | 0.11 | 2.9E-07 | 4.5E-03 |
| **Trait / Analysis: Diffuse opacities (quantitative) / p-difference (Sex stratified)** | | | | | | |
| GO_cc:go_alpha_beta_t_cell_receptor_complex | 5 | 1.6 | 0.03 | 0.33 | 3.9E-07 | 6.0E-03 |
| **Trait / Analysis: Hypoplastic defects (Binary) / Main analysis** | | | | | | |
| GO_bp:go_elastic_fiber_assembly | 8 | 1.1 | 0.02 | 0.24 | 1.2E-06 | 1.9E-02 |
| **Trait / Analysis: Hypoplastic defects (Binary) / Female-specific GWAS** | | | | | | |
| GO_cc:go_stereocilium_membrane | 4 | 1.8 | 0.03 | 0.38 | 8.4E-07 | 1.3E-02 |

**Abbreviations:** p-diff: p-value for the difference of the stratum-specific beta coefficients; SE: Standard Error; P_bon_: Bonferroni corrected p-value; Beta: Regression coefficient; Beta STD: Standardized regression coefficient

**Appendix Table 10.** Enrichment of prioritized genes in curated genesets (Reporting gene sets with adjusted P-value< 5 x10^-8^)

| **Category** | **GeneSet** | **N_genes** | **N_overlap** | **p** | **adjP*** | ***genes*** |
| --- | --- | --- | --- | --- | --- | --- |
| **Trait / Analysis: Demarcated opacities (Quantitative) - Main analysis** | | | | | | |
| Positional gene sets | chr20q13 | 225 | 31 | 9.3E-37 | 2.8E-34 | *TSHZ2, ADRM1, LAMA5, C20orf166, SLCO4A1, NTSR1, MRGBP, OGFR, COL9A3, DIDO1, GID8, SLC17A9, YTHDF1, ARFGAP1, COL20A1, KCNQ2, STMN3, RTEL1, RTEL1-TNFRSF6B, TNFRSF6B, ARFRP1, ZGPAT, LIME1, SLC2A4RG, ZBTB46, DNAJC5, UCKL1, ZNF512B, PRPF6, TCEA2, RGS19* |
| Curated_gene_sets | NIKOLSKY_BREAST_CANCER_20Q12_Q13_AMPLICON | 146 | 30 | 4.0E-41 | 2.2E-37 | *TSHZ2, ADRM1, LAMA5, C20orf166, SLCO4A1, NTSR1, MRGBP, OGFR, COL9A3, DIDO1, GID8, SLC17A9, YTHDF1, ARFGAP1, COL20A1, KCNQ2, STMN3, RTEL1, TNFRSF6B, ARFRP1, ZGPAT, LIME1, SLC2A4RG, ZBTB46, DNAJC5, UCKL1, ZNF512B, PRPF6, TCEA2, RGS19* |
| Chemical and Genetic pertubation gene sets | NIKOLSKY_BREAST_CANCER_20Q12_Q13_AMPLICON | 146 | 30 | 4.0E-41 | 1.3E-37 | *TSHZ2, ADRM1, LAMA5, C20orf166, SLCO4A1, NTSR1, MRGBP, OGFR, COL9A3, DIDO1, GID8, SLC17A9, YTHDF1, ARFGAP1, COL20A1, KCNQ2, STMN3, RTEL1, TNFRSF6B, ARFRP1, ZGPAT, LIME1, SLC2A4RG, ZBTB46, DNAJC5, UCKL1, ZNF512B, PRPF6, TCEA2, RGS19* |
| GWAS catalog reported genes | Prostate cancer | 302 | 17 | 1.3E-13 | 2.3E-10 | *PPFIBP2, ADRM1, LAMA5, C20orf166, STMN3, RTEL1, RTEL1-TNFRSF6B, TNFRSF6B, ARFRP1, ZGPAT, LIME1, SLC2A4RG, ZBTB46, DNAJC5, UCKL1, ZNF512B, PRPF6* |
| **Trait / Analysis: Diffuse opacities (Quantitative) - Main analysis** | | | | | | |
| Positional gene sets | chr12q13 | 265 | 11 | 1.1E-11 | 3.4E-09 | *ARHGAP9, MARS, DDIT3, MBD6, DCTN2, KIF5A, PIP4K2C, DTX3, ARHGEF25, SLC26A10, B4GALNT1* |
| **Trait / Analysis: Hypoplastic defects (Quantitative) - Main analysis** | | | | | | |
| GWAS catalog reported genes | Facial emotion recognition (sad faces) | 23 | 10 | 1.9E-15 | 3.5E-12 | *ARPC1A, ARPC1B, PDAP1, BUD31, PTCD1, CPSF4, ATP5J2, ZNF789, ZKSCAN5, FAM200A* |
| **Trait / Analysis: Demarcated opacities (Quantitative) / Joint test - Sex-stratified** | | | | | | |
| Positional gene sets | chr20q13 | 225 | 31 | 1.5E-32 | 4.4E-30 | *TSHZ2, ADRM1, LAMA5, C20orf166, SLCO4A1, NTSR1, MRGBP, OGFR, COL9A3, DIDO1, GID8, SLC17A9, YTHDF1, ARFGAP1, COL20A1, KCNQ2, STMN3, RTEL1, RTEL1-TNFRSF6B, TNFRSF6B, ARFRP1, ZGPAT, LIME1, SLC2A4RG, ZBTB46, DNAJC5, UCKL1, ZNF512B, PRPF6, TCEA2, RGS19* |
| Curated_gene_sets | NIKOLSKY_BREAST_CANCER_20Q12_Q13_AMPLICON | 146 | 30 | 4.8E-37 | 2.7E-33 | *TSHZ2, ADRM1, LAMA5, C20orf166, SLCO4A1, NTSR1, MRGBP, OGFR, COL9A3, DIDO1, GID8, SLC17A9, YTHDF1, ARFGAP1, COL20A1, KCNQ2, STMN3, RTEL1, TNFRSF6B, ARFRP1, ZGPAT, LIME1, SLC2A4RG, ZBTB46, DNAJC5, UCKL1, ZNF512B, PRPF6, TCEA2, RGS19* |
| Chemical and Genetic pertubation gene sets | NIKOLSKY_BREAST_CANCER_20Q12_Q13_AMPLICON | 146 | 30 | 4.8E-37 | 1.6E-33 | *TSHZ2, ADRM1, LAMA5, C20orf166, SLCO4A1, NTSR1, MRGBP, OGFR, COL9A3, DIDO1, GID8, SLC17A9, YTHDF1, ARFGAP1, COL20A1, KCNQ2, STMN3, RTEL1, TNFRSF6B, ARFRP1, ZGPAT, LIME1, SLC2A4RG, ZBTB46, DNAJC5, UCKL1, ZNF512B, PRPF6, TCEA2, RGS19* |
| GWAS catalog reported genes | Alcohol consumption (max-drinks) | 17 | 8 | 6.1E-14 | 1.1E-10 | *FRMD4A, BRAP, ACAD10, NAA25, HECTD4, PTPN11, RPH3A, TSHZ2* |
|  | Prostate cancer | 302 | 17 | 1.3E-11 | 1.2E-08 | *PPFIBP2, ADRM1, LAMA5, C20orf166, STMN3, RTEL1, RTEL1-TNFRSF6B, TNFRSF6B, ARFRP1, ZGPAT, LIME1, SLC2A4RG, ZBTB46, DNAJC5, UCKL1, ZNF512B, PRPF6* |
| **Trait / Analysis: Demarcated opacities (Quantitative) / Male-specific GWAS** | | | | | | |
| Positional gene sets | chr20q13 | 225 | 29 | 1.7E-30 | 5.1E-28 | *ADRM1, LAMA5, C20orf166, NTSR1, MRGBP, OGFR, COL9A3, DIDO1, GID8, SLC17A9, YTHDF1, ARFGAP1, COL20A1, KCNQ2, STMN3, RTEL1, RTEL1-TNFRSF6B, TNFRSF6B, ARFRP1, ZGPAT, LIME1, SLC2A4RG, ZBTB46, DNAJC5, UCKL1, ZNF512B, PRPF6, TCEA2, RGS19* |
| Curated_gene_sets | NIKOLSKY_BREAST_CANCER_20Q12_Q13_AMPLICON | 146 | 28 | 1.5E-34 | 8.3E-31 | *ADRM1, LAMA5, C20orf166, NTSR1, MRGBP, OGFR, COL9A3, DIDO1, GID8, SLC17A9, YTHDF1, ARFGAP1, COL20A1, KCNQ2, STMN3, RTEL1, TNFRSF6B, ARFRP1, ZGPAT, LIME1, SLC2A4RG, ZBTB46, DNAJC5, UCKL1, ZNF512B, PRPF6, TCEA2, RGS19* |
| Chemical and Genetic pertubation gene sets | NIKOLSKY_BREAST_CANCER_20Q12_Q13_AMPLICON | 146 | 28 | 1.5E-34 | 5.0E-31 | *ADRM1, LAMA5, C20orf166, NTSR1, MRGBP, OGFR, COL9A3, DIDO1, GID8, SLC17A9, YTHDF1, ARFGAP1, COL20A1, KCNQ2, STMN3, RTEL1, TNFRSF6B, ARFRP1, ZGPAT, LIME1, SLC2A4RG, ZBTB46, DNAJC5, UCKL1, ZNF512B, PRPF6, TCEA2, RGS19* |
| GWAS catalog reported genes | Alcohol consumption (max-drinks) | 17 | 7 | 4.9E-12 | 8.9E-09 | *FRMD4A, BRAP, ACAD10, NAA25, HECTD4, PTPN11, RPH3A* |
|  | Prostate cancer | 302 | 16 | 4.8E-11 | 4.3E-08 | *ADRM1, LAMA5, C20orf166, STMN3, RTEL1, RTEL1-TNFRSF6B, TNFRSF6B, ARFRP1, ZGPAT, LIME1, SLC2A4RG, ZBTB46, DNAJC5, UCKL1, ZNF512B, PRPF6* |
| **Trait / Analysis: Developmental defects of enamel (Quantitative) / P-difference (Sex-stratified)** | | | | | | |
| Positional gene sets | chr5q31 | 186 | 11 | 1.6E-15 | 4.8E-13 | *PCDHB5, PCDHB6, PCDHB17, PCDHB7, PCDHB8, PCDHB16, PCDHB10, PCDHB11, PCDHB12, PCDHB13, PCDHB14* |
| GO biological processes | GO_HOMOPHILIC_CELL_ADHESION_VIA_PLASMA_MEMBRANE_ADHESION_MOLECULES | 163 | 11 | 3.7E-16 | 2.7E-12 | *CDH10, PCDHB5, PCDHB6, PCDHB7, PCDHB8, PCDHB16, PCDHB10, PCDHB11, PCDHB12, PCDHB13, PCDHB14* |
|  | GO_CELL_CELL_ADHESION_VIA_PLASMA_MEMBRANE_ADHESION_MOLECULES | 267 | 12 | 1.8E-15 | 6.5E-12 | *CDH10, PCDHB5, PCDHB6, PCDHB7, PCDHB8, PCDHB16, PCDHB10, PCDHB11, PCDHB12, PCDHB13, PCDHB14, TENM2* |
|  | GO_CALCIUM_DEPENDENT_CELL_CELL_ADHESION_VIA_PLASMA_MEMBRANE_CELL_ADHESION_MOLECULES | 47 | 8 | 2.9E-15 | 7.1E-12 | *CDH10, PCDHB5, PCDHB6, PCDHB16, PCDHB10, PCDHB11, PCDHB13, PCDHB14* |
| GO molecular functions | GO_CALCIUM_ION_BINDING | 680 | 13 | 5.2E-12 | 8.5E-09 | *NECAB2, CDH10, PCDHB5, PCDHB6, PCDHB7, PCDHB8, PCDHB16, PCDHB10, PCDHB11, PCDHB12, PCDHB13, PCDHB14, TENM2* |

**Appendix Table 11.** Generalization of loci discovered using Approach 1 in the ZOE-2.0 study to summary statistics from Alotaibi et al. 2023 (PMID: 35172313).

| **Zoe 2.0 study** | | | | | | **Alotaibi 2023** | | | |
| --- | --- | --- | --- | --- | --- | --- | --- | --- | --- |
| **Locus** | **rsid** | **SNPs (LD R^2^>0.80)** | **P** | **Beta** | **P** | | **Beta** | | **Directional consistency** |
| **Developmental defects of enamel (Quantitative)** | | | | | |  | |  | |
| *Y_RNA* | rs143210152 |  | 4.4E-09 | 0.45 | na | | na | |  |
|  |  | rs150884264 |  |  | na | | na | |  |
|  |  | rs78792587 |  |  | na | | na | |  |
|  |  | rs74363957 |  |  | na | | na | |  |
|  |  | rs77669438 |  |  | na | | na | |  |
| *DEK* | rs567243779 |  | 8.1E-09 | 0.23 | na | | na | |  |
|  |  | rs12189968 |  |  | 2.5E-01 | | -0.011 | | No |
|  |  | rs12200177 |  |  | 2.5E-01 | | -0.011 | | No |
| *RP4-724E13.2* | rs150075871 |  | 2.0E-08 | 0.53 | na | | na | |  |
| *HBS1L* | rs116989668 |  | 4.0E-08 | 0.52 | na | | na | |  |
| **Demarcated opacities (Quantitative)** | | | | | |  | |  | |
| *SLCO4A1:RP11-93B14.5* | rs75880235 |  | 1.9E-08 | 0.51 | na | | na | |  |
| *C21orf54* | rs895438538 |  | 4.2E-08 | 0.33 | na | | na | |  |
| **Diffuse opacities (Quantitative)** | | | | | |  | |  | |
| *Y_RNA* | rs77669438 |  | 9.1E-11 | 0.56 | na | | na | |  |
|  |  | rs74363957 |  |  | na | | na | |  |
|  |  | rs143210152 |  |  | na | | na | |  |
|  |  | rs150884264 |  |  | na | | na | |  |
|  |  | rs78792587 |  |  | na | | na | |  |
| *CTA-21C21.1* | rs1658697761 |  | 2.0E-09 | 0.11 | na | | na | |  |
|  |  | rs4656024 |  |  | 8.3E-01 | | -0.001 | | No |
|  |  | rs1889956 |  |  | 8.2E-01 | | -0.001 | | No |
|  |  | rs10801880 |  |  | 3.4E-01 | | -0.005 | | No |
|  |  | rs1932759 |  |  | 3.7E-01 | | -0.005 | | No |
|  |  | rs12133011 |  |  | 3.9E-01 | | -0.005 | | No |
| **Hypoplastic defects (Quantitative)** | | | | | |  | |  | |
| *RP11-157E21.2* | rs192067416 |  | 2.4E-10 | 0.46 | na | | na | |  |
|  |  | rs75243303 |  |  | na | | na | |  |
|  |  | rs79969289 |  |  | na | | na | |  |
| *AC007950.1* | rs79505735 |  | 5.1E-10 | 0.46 | na | | na | |  |
| *RP1-167G20.1* | rs149507176 |  | 1.5E-09 | 0.52 | na | | na | |  |
|  |  | rs116554553 |  |  | na | | na | |  |
|  |  | rs146957249 |  |  | na | | na | |  |
|  |  | rs115597729 |  |  | na | | na | |  |
|  |  | rs145692407 |  |  | na | | na | |  |
|  |  | rs180734684 |  |  | na | | na | |  |
| *ADAMTSL1* | rs115373518 |  | 7.5E-09 | 0.32 | 7.9E-01 | | 0.014 | | Yes |
| *RAVER2* | -- |  | 7.6E-09 | 0.50 | na | | na | |  |
| *FHIT* | rs73835238 |  | 9.1E-09 | 0.52 | na | | na | |  |
| *UBE2G2* | rs115617269 |  | 2.0E-08 | 0.43 | 6.6E-01 | | 0.020 | | Yes |
|  |  | rs115478160 |  |  | na | | na | |  |
|  |  | rs114237344 |  |  | na | | na | |  |
| *MPV17L2* | rs79028494 |  | 2.9E-08 | 0.28 | **3.2E-02** | | -0.021 | | No |
|  |  | rs149848864 |  |  | na | | na | |  |
|  |  | rs115420983 |  |  | **3.2E-02** | | -0.021 | | No |
|  |  | rs143418651 |  |  | na | | na | |  |
|  |  | rs12150905 |  |  | **3.2E-02** | | -0.021 | | No |
|  |  | rs11552160 |  |  | **2.5E-02** | | -0.023 | | No |
|  |  | rs141596690 |  |  | na | | na | |  |
|  |  | rs148340002 |  |  | na | | na | |  |
|  |  | rs116893986 |  |  | **3.2E-02** | | -0.021 | | No |
|  |  | rs115424305 |  |  | **3.2E-02** | | -0.021 | | No |
| *PPAPDC3* | rs75061854 |  | 2.9E-08 | 0.41 | NA | | na | |  |
| *CLUL1* | rs114989924 |  | 3.6E-08 | 0.37 | na | | na | |  |
| *MAML2* | rs150489138 |  | 4.3E-08 | 0.45 | na | | na | |  |
|  |  | rs186550701 |  |  | na | | na | |  |
| *MTHFD1L* | rs11963290 |  | 4.8E-08 | 0.17 | 7.9E-01 | | -0.005 | | No |
| **Diffuse opacities (Binary)** | | | | | |  | |  | |
| *RP1-281H8.3* | rs184135739 |  | 3.0E-09 | 0.98 | na | | na | |  |
|  |  | rs182094308 |  |  | na | | na | |  |
|  |  | rs192636896 |  |  | na | | na | |  |

Nominally significant (P<0.05)

Bonferroni-corrected significance (P<1.3x10^-3^)

**Appendix Table 12.** Generalization of loci discovered using Approach 2-3 in the ZOE-2.0 study to summary statistics from Alotaibi et al. 2023 (PMID: 35172313).

| **ZOE-2.0 study** | | | | | | **Alotaibi 2023** | | | |
| --- | --- | --- | --- | --- | --- | --- | --- | --- | --- |
| **Locus name** | **rsid** | **SNPs (LD R^2^>0.8)** | **P** | **Beta** | **P** | | **Beta** | | **Directional consistency** |
| **Quantitative traits** | | | | | |  | |  | |
| **Developmental defects of enamel - Female stratum** | | | | | |  | |  | |
| *ALDH1A1* | rs141753205 |  | 9.6E-10 | 0.58 | na | | na | |  |
| *RP11-384F7.1* | rs202021524 |  | 4.8E-08 | 0.51 | na | | na | |  |
| **Developmental defects of enamel - Male stratum** | | | | | |  | |  | |
| *RP11-415K20.1* | rs16826979 |  | 8.4E-09 | 0.63 | na | | na | |  |
|  |  | rs144782318 |  |  | na | | na | |  |
| *TMPRSS13* | rs117786109 |  | 4.8E-08 | 0.49 | 8.7E-01 | | -0.005 | | No |
| **Developmental defects of enamel - Joint 2df test** | | | | | |  | |  | |
| *Y_RNA* | rs143210152 |  | 2.1E-08 | - | na | | na | |  |
|  |  | rs150884264 |  |  | na | | na | |  |
|  |  | rs78792587 |  |  | na | | na | |  |
|  |  | rs74363957 |  |  | na | | na | |  |
|  |  | rs77669438 |  |  | na | | na | |  |
| **Demarcated opacities - Female stratum** | | | | | |  | |  | |
| *JRKL:JRKL-AS1* | rs141989840 |  | 1.1E-08 | 0.49 | na | | na | |  |
|  |  | rs115852593 |  |  | 7.9E-01 | | 0.015 | | Yes |
|  |  | rs7948994 |  |  | 7.9E-01 | | 0.015 | | Yes |
|  |  | rs149346861 |  |  | na | | na | |  |
|  |  | rs142336354 |  |  | na | | na | |  |
|  |  | rs76402869 |  |  | 7.3E-01 | | 0.017 | | Yes |
|  |  | rs114323294 |  |  | na | | na | |  |
|  |  | rs146164526 |  |  | na | | na | |  |
|  |  | rs141422323 |  |  | na | | na | |  |
|  |  | rs115503775 |  |  | 7.3E-01 | | 0.017 | | Yes |
|  |  | rs142954847 |  |  | na | | na | |  |
|  |  | rs74370899 |  |  | 7.3E-01 | | 0.017 | | Yes |
|  |  | rs141098155 |  |  | na | | na | |  |
|  |  | rs147734273 |  |  | na | | na | |  |
|  |  | rs554439578 |  |  | na | | na | |  |
|  |  | rs7126970 |  |  | 7.3E-01 | | 0.017 | | Yes |
|  |  | rs7126813 |  |  | 7.3E-01 | | 0.017 | | Yes |
|  |  | rs183940017 |  |  | na | | na | |  |
|  |  | rs140973057 |  |  | na | | na | |  |
|  |  | rs115290383 |  |  | 2.5E-01 | | -0.050 | | No |
|  |  | rs7928839 |  |  | 7.3E-01 | | 0.017 | | Yes |
|  |  | rs143175562 |  |  | na | | na | |  |
|  |  | rs115197601 |  |  | 7.3E-01 | | 0.017 | | Yes |
|  |  | rs115384750 |  |  | 7.3E-01 | | 0.017 | | Yes |
|  |  | rs143823963 |  |  | na | | na | |  |
|  |  | rs114196266 |  |  | 7.3E-01 | | 0.017 | | Yes |
|  |  | rs115314907 |  |  | 7.3E-01 | | 0.017 | | Yes |
|  |  | rs116604368 |  |  | 7.3E-01 | | 0.017 | | Yes |
|  |  | rs191599231 |  |  | na | | na | |  |
|  |  | rs116794369 |  |  | 7.4E-01 | | 0.016 | | Yes |
| *RP11-534P19.1* | rs61471572 |  | 2.1E-08 | 0.63 | na | | na | |  |
|  |  | rs146237485 |  |  | na | | na | |  |
|  |  | rs59268482 |  |  | na | | na | |  |
|  |  | rs57691714 |  |  | na | | na | |  |
|  |  | rs61040565 |  |  | na | | na | |  |
|  |  | rs59292726 |  |  | na | | na | |  |
| *DLGAP2* | rs2906574 |  | 3.4E-08 | -0.53 | 8.1E-02 | | 0.022 | | No |
|  |  | rs2906575 |  |  | **2.2E-02** | | 0.081 | | No |
|  |  | rs2906589 |  |  | 7.8E-02 | | 0.022 | | No |
| *FHIT* | rs72891211 |  | 4.4E-08 | 0.62 | **3.1E-03** | | -0.122 | | No |
|  |  | rs111642535 |  |  | **3.1E-03** | | -0.122 | | No |
|  |  | rs72889167 |  |  | **3.1E-03** | | -0.122 | | No |
|  |  | rs113302649 |  |  | **3.1E-03** | | -0.123 | | No |
| **Demarcated opacities - Male stratum** | | | | | |  | |  | |
| *PTPRK* | rs77758675 |  | 7.0E-10 | 0.74 | 7.4E-01 | | 0.008 | | Yes |
|  |  | rs75451681 |  |  | 7.5E-01 | | 0.007 | | Yes |
|  |  | rs78677308 |  |  | 7.6E-01 | | 0.007 | | Yes |
|  |  | rs117058546 |  |  | 7.4E-01 | | 0.008 | | Yes |
|  |  | rs117509604 |  |  | 7.7E-01 | | 0.007 | | Yes |
| *RERE* | rs77921067 |  | 1.5E-09 | 0.70 | na | | na | |  |
|  |  | rs187053242 |  |  | na | | na | |  |
|  |  | rs144935761 |  |  | na | | na | |  |
| *ZNRF3* | rs369163187 |  | 3.1E-09 | 0.68 | na | | na | |  |
| *RP11-57H14.3* | rs77466522 |  | 3.2E-09 | 0.58 | 8.8E-01 | | -0.005 | | No |
| *EFR3B* | rs188016617 |  | 9.9E-09 | 0.73 | na | | na | |  |
| *FRMD5* | rs146484655 |  | 3.7E-08 | 0.68 | na | | na | |  |
|  |  | rs146484655 |  |  | na | | na | |  |
|  |  | rs75668125 |  |  | na | | na | |  |
|  |  | rs147244518 |  |  | na | | na | |  |
|  |  | rs140645911 |  |  | na | | na | |  |
|  |  | rs574411972 |  |  | na | | na | |  |
|  |  | rs145438657 |  |  | na | | na | |  |
|  |  | rs115206077 |  |  | na | | na | |  |
|  |  | rs114710495 |  |  | na | | na | |  |
|  |  | rs143156550 |  |  | na | | na | |  |
|  |  | rs79259027 |  |  | na | | na | |  |
| *RP5-963E22.4* | rs201297757 |  | 3.7E-08 | 0.62 | na | | na | |  |
| *LINC01508* | rs569532598 |  | 4.6E-08 | 0.72 | na | | na | |  |
|  |  | rs199781599 |  |  | na | | na | |  |
| **Demarcated opacities - p-diff** | | | | | |  | |  | |
| *SLC2A3* | rs61921622 |  | 1.8E-08 | - | 3.2E-01 | | 0.012 | | na |
| **Diffuse opacities - Female stratum** | | | | | |  | |  | |
| *GBE1* | rs57583981 |  | 4.5E-09 | 0.59 | 9.1E-01 | | 0.019 | | Yes |
|  |  | rs149085411 |  |  | na | | na | |  |
|  |  | rs3772892 |  |  | na | | na | |  |
|  |  | rs118172718 |  |  | **1.4E-03** | | -0.298 | | No |
|  |  | rs139962089 |  |  | na | | na | |  |
| *TENM2* | rs141872488 |  | 1.6E-08 | 0.58 | na | | na | |  |
| *THBS4* | rs77790627 |  | 2.1E-08 | 0.59 | 9.1E-01 | | 0.018 | | Yes |
| *RP11-384F7.1* | -- |  | 3.8E-08 | 0.51 | na | | na | |  |
| **Diffuse opacities - Male stratum** | | | | | |  | |  | |
| *RP11-473I1.6* | rs140095098 |  | 2.9E-08 | 0.78 | na | | na | |  |
| **Diffuse opacities - Joint 2df test** | | | | | |  | |  | |
| *Y_RNA* | rs77669438 |  | 1.3E-09 | - | na | | na | |  |
|  |  | rs74363957 |  |  | na | | na | |  |
|  |  | rs143210152 |  |  | na | | na | |  |
|  |  | rs150884264 |  |  | na | | na | |  |
|  |  | rs78792587 |  |  | na | | na | |  |
| *CTA-21C21.1* | rs11440629 | Not in 1000G panel | 1.5E-08 | - | na | | na | |  |
| **Hypoplastic defects - Female stratum** | | | | | |  | |  | |
| *ADAMTSL1* | rs55947174 |  | 1.5E-11 | 0.74 | 9.2E-01 | | 0.011 | | Yes |
| *KRTAP7-1* | rs111896551 |  | 5.4E-10 | 0.59 | na | | na | |  |
| *ADD2* | rs146459483 |  | 8.3E-10 | 0.75 | na | | na | |  |
| *CNTN4* | -- |  | 1.6E-09 | 0.83 | na | | na | |  |
| *STARP1* | rs973279911 |  | 1.9E-09 | 0.74 | na | | na | |  |
| *HPD* | rs568971993 |  | 2.0E-09 | 0.70 | na | | na | |  |
|  |  | rs118055116 |  |  | 5.4E-01 | | -0.009 | | No |
|  |  | rs141976264 |  |  | na | | na | |  |
|  |  | rs184954033 |  |  | na | | na | |  |
|  |  | rs140552304 |  |  | na | | na | |  |
|  |  | rs78258889 |  |  | 3.7E-01 | | -0.013 | | No |
| *TMEM40* | rs180978195 |  | 2.4E-09 | 0.87 | na | | na | |  |
| *GABRG3* | rs191103614 |  | 5.5E-09 | 0.82 | na | | na | |  |
|  |  | rs182450425 |  |  | na | | na | |  |
| *CCSER1* | rs139393106 |  | 8.0E-09 | 0.55 | na | | na | |  |
|  |  | rs144071308 |  |  | na | | na | |  |
|  |  | rs182744873 |  |  | na | | na | |  |
|  |  | rs77443826 |  |  | 8.6E-01 | | 0.028 | | Yes |
| *ZNF331* | rs192307826 |  | 1.3E-08 | 0.75 | na | | na | |  |
| *JRKL:JRKL-AS1* | rs11826357 |  | 1.4E-08 | 0.72 | na | | na | |  |
| *MMP26* | rs17251629 |  | 1.6E-08 | 0.76 | 6.9E-01 | | -0.013 | | No |
| *CTD-2021A8.3* | rs140671047 |  | 1.6E-08 | 0.73 | na | | na | |  |
| *MCTP2* | rs117304168 |  | 1.6E-08 | 0.72 | na | | na | |  |
| *ZNF823* | -- |  | 1.7E-08 | 0.78 | na | | na | |  |
| *CYP4F22* | rs149762022 |  | 2.0E-08 | 0.69 | na | | na | |  |
|  |  | rs186101958 |  |  | na | | na | |  |
| *PLEKHD1* | rs147589787 |  | 2.3E-08 | 0.56 | na | | na | |  |
| *MAST4* | rs1753671813 |  | 4.5E-08 | 0.55 | na | | na | |  |
|  |  | rs11323919 |  |  | na | | na | |  |
|  |  | rs529117682 |  |  | na | | na | |  |
|  |  | rs114926884 |  |  | na | | na | |  |
|  |  | rs76572286 |  |  | na | | na | |  |
|  |  | rs115181284 |  |  | na | | na | |  |
|  |  | rs116578009 |  |  | na | | na | |  |
|  |  | rs116835797 |  |  | na | | na | |  |
|  |  | rs115580712 |  |  | na | | na | |  |
|  |  | rs115155623 |  |  | na | | na | |  |
|  |  | rs115307115 |  |  | na | | na | |  |
|  |  | rs150352837 |  |  | na | | na | |  |
|  |  | rs114963452 |  |  | na | | na | |  |
|  |  | rs116577066 |  |  | na | | na | |  |
|  |  | rs115613284 |  |  | na | | na | |  |
|  |  | rs57088072 |  |  | na | | na | |  |
| *NF1* | rs147836184 |  | 4.6E-08 | 0.58 | na | | na | |  |
|  |  | rs59458028 |  |  | na | | na | |  |
|  |  | rs114923762 |  |  | na | | na | |  |
| **Hypoplastic defects - Male stratum** | | | | | |  | |  | |
| *AC073928.2* | rs141261078 |  | 4.7E-10 | 0.71 | na | | na | |  |
| *GADL1* | rs144577905 |  | 2.1E-09 | 0.72 | na | | na | |  |
| *PDCD1LG2* | rs138482896 |  | 2.3E-09 | 0.80 | na | | na | |  |
|  |  | rs138857354 |  |  | na | | na | |  |
| *RP11-586E1.1* | rs115583696 |  | 5.1E-09 | 0.71 | na | | na | |  |
| *RAVER2* | -- |  | 6.7E-09 | 0.73 | na | | na | |  |
| *PIK3R4* | rs150504962 | Not in 1000G panel | 7.4E-09 | 0.66 | na | | na | |  |
| *CCDC18* | rs187188755 |  | 2.4E-08 | 0.55 | na | | na | |  |
| *BCHE* | rs6805552 |  | 3.2E-08 | -0.63 | 7.4E-01 | | -0.006 | | Yes |
| *AGTRAP* | rs118165546 |  | 3.4E-08 | 0.77 | na | | na | |  |
|  |  | rs138499116 |  |  | na | | na | |  |
|  |  | rs59176666 |  |  | na | | na | |  |
|  |  | rs78895176 |  |  | na | | na | |  |
|  |  | rs76125441 |  |  | na | | na | |  |
|  |  | rs142645237 |  |  | na | | na | |  |
|  |  | rs141361257 |  |  | na | | na | |  |
|  |  | rs143863509 |  |  | na | | na | |  |
|  |  | rs148618845 |  |  | na | | na | |  |
|  |  | rs117093171 |  |  | na | | na | |  |
| *KRTAP9-6* | rs2016470489 | Not in 1000G panel | 3.7E-08 | 0.94 | na | | na | |  |
| *WI2-81516E3.1* | rs146461752 |  | 4.5E-08 | 0.65 | na | | na | |  |
|  |  | rs9628066 |  |  | na | | na | |  |
| *TENM2* | rs62383285 |  | 4.9E-08 | 0.56 | 8.0E-01 | | -0.008 | | No |
| **Hypoplastic defects - Joint 2df test** | | | | | |  | |  | |
| *AC007950.1* | rs79505735 |  | 8.1E-09 | - | na | | na | |  |
| *RP1-167G20.1* | rs149507176 |  | 1.2E-08 | - | na | | na | |  |
|  |  | rs116554553 |  |  | na | | na | |  |
|  |  | rs146957249 |  |  | na | | na | |  |
|  |  | rs115597729 |  |  | na | | na | |  |
|  |  | rs145692407 |  |  | na | | na | |  |
|  |  | rs180734684 |  |  | na | | na | |  |
| *RP11-157E21.2* | rs192067416 |  | 1.3E-08 | - | na | | na | |  |
|  |  | rs75243303 |  |  | na | | na | |  |
|  |  | rs79969289 |  |  | na | | na | |  |
| *CCDC82* | rs192993105 |  | 4.2E-08 | - | na | | na | |  |
|  |  | rs11826357 |  |  | na | | na | |  |
|  |  | rs144780667 |  |  | na | | na | |  |
|  |  | rs185056538 |  |  | na | | na | |  |
|  |  | rs115970058 |  |  | na | | na | |  |
|  |  | rs144787154 |  |  | na | | na | |  |
| **Binary traits** | | | | | |  | |  | |
| **Developmental defects of enamel - Male stratum** | | | | | |  | |  | |
| *SLC19A1* | rs62214342 |  | 3.9E-08 | 0.34 | 4.0E-01 | | -0.005 | | No |
|  |  | rs57044206 |  |  | 3.2E-01 | | -0.006 | | No |
|  |  | rs8129273 |  |  | 3.1E-01 | | -0.006 | | No |
|  |  | rs7349007 |  |  | 2.9E-01 | | -0.006 | | No |
|  |  | rs58240688 |  |  | 2.6E-01 | | -0.006 | | No |
|  |  | rs75025769 |  |  | 2.6E-01 | | -0.006 | | No |
| **Developmental defects of enamel - p-diff** | | | | | |  | |  | |
| *snoU13* | rs7228955 |  | 4.0E-08 | - | 7.7E-01 | | -0.002 | | na |
|  |  | rs4488566 |  |  | 6.9E-01 | | -0.002 | | na |
| **Demarcated opacities - Male stratum** | | | | | |  | |  | |
| *CTD-2644I21.1* | rs2776544 |  | 3.2E-09 | 0.34 | 2.2E-01 | | 0.007 | | Yes |
| *RP11-203H2.1* | rs17143504 |  | 3.9E-08 | -0.44 | 7.0E-01 | | -0.003 | | Yes |
|  |  | rs62398162 |  |  | 6.2E-01 | | -0.004 | | Yes |
|  |  | rs17143510 |  |  | 6.2E-01 | | -0.004 | | Yes |
|  |  | rs62398164 |  |  | 6.5E-01 | | -0.004 | | Yes |
| **Diffuse opacities - Male stratum** | | | | | |  | |  | |
| *LINC00670* | rs11078084 |  | 2.1E-08 | -0.29 | 7.1E-01 | | -0.002 | | Yes |
|  |  | rs11078084 |  |  | 7.1E-01 | | -0.002 | | Yes |
|  |  | rs56084989 |  |  | na | | na | |  |
|  |  | rs35790736 |  |  | na | | na | |  |
|  |  | rs7221414 |  |  | na | | na | |  |
|  |  | rs60459111 |  |  | na | | na | |  |
|  |  | rs9901558 |  |  | na | | na | |  |
|  |  | rs9901652 |  |  | na | | na | |  |
|  |  | rs12453228 |  |  | na | | na | |  |
|  |  | rs12453235 |  |  | na | | na | |  |
|  |  | rs11078086 |  |  | na | | na | |  |
|  |  | rs1519249 |  |  | na | | na | |  |
|  |  | rs111311447 |  |  | na | | na | |  |
|  |  | rs55977007 |  |  | na | | na | |  |
|  |  | rs1519250 |  |  | na | | na | |  |
|  |  | rs7217115 |  |  | na | | na | |  |
|  |  | rs12450472 |  |  | na | | na | |  |
|  |  | rs1989503 |  |  | na | | na | |  |
|  |  | rs1074721 |  |  | na | | na | |  |
|  |  | rs148380282 |  |  | na | | na | |  |
|  |  | rs1961550 |  |  | na | | na | |  |
|  |  | rs1989504 |  |  | na | | na | |  |
|  |  | rs6502197 |  |  | na | | na | |  |
|  |  | rs1074724 |  |  | na | | na | |  |
|  |  | rs1544510 |  |  | 3.3E-01 | | 0.006 | | No |
|  |  | rs1989505 |  |  | na | | na | |  |
|  |  | rs1544509 |  |  | na | | na | |  |
|  |  | rs12601969 |  |  | na | | na | |  |
|  |  | rs4054988 |  |  | na | | na | |  |
|  |  | rs7224880 |  |  | na | | na | |  |
|  |  | rs1074720 |  |  | na | | na | |  |

Nominally significant (P<0.05)

Bonferroni-corrected significance (P<1.3x10^-3^)

**Appendix Table 13.** Generalization of loci identified in the main discovery GWAS (Approach 1) to GLIDE children and GLIDE adult results.

| Locus Name | rsid | Chr:Pos(hg19) | | EA/OA | | Beta | | GLIDE | | GLIDE children (Primary teeth_multiethnic) | | | | | | | | GLIDE children (Permanent teeth) | | | | | | | | GLIDE adults (DMFS and Dentures traits) | | | | | | |
| --- | --- | --- | --- | --- | --- | --- | --- | --- | --- | --- | --- | --- | --- | --- | --- | --- | --- | --- | --- | --- | --- | --- | --- | --- | --- | --- | --- | --- | --- | --- | --- | --- |
|  |  |  | |  | |  | | **EA/OA** | | **Beta** | | **P** | | **N** | | **DC** | | **Beta** | | **P** | | **N** | | **DC** | | **Beta** | | **P** | | **Effective N** | | **DC** |
| Developmental defects of enamel (quantitative) | | | | |  | |  | |  | |  | |  | |  | |  | |  | |  | |  | |  | |  | |  | |  | |
| Y_RNA | rs143210152 | chr7:155957044 | | G/C | | 0.98 | |  | |  | |  | |  | |  | |  | |  | |  | |  | |  | |  | |  | |  |
| DEK | rs567243779 | chr6:18228721 | | A/C | | 0.23 | |  | |  | |  | |  | |  | |  | |  | |  | |  | |  | |  | |  | |  |
|  | rs12189968** | chr6:18216591 | |  | |  | | t/c | | -0.02 | | 6.4E-01 | | 18999 | | No | | 0.04 | | 4.3E-01 | | 13385 | | Yes | | 0.00 | | 9.6E-01 | | 285247 | | Yes |
| RP4-724E13.2 | rs150075871 | chr7:50947954 | | T/C | | 0.98 | |  | |  | |  | |  | |  | |  | |  | |  | |  | |  | |  | |  | |  |
| HBS1L | rs116989668 | chr6:135334350 | | A/G | | 0.52 | |  | |  | |  | |  | |  | |  | |  | |  | |  | |  | |  | |  | |  |
| Demarcated opacities (quantitative) | | |  | |  | |  | |  | |  | |  | |  | |  | |  | |  | |  | |  | |  | |  | |  | |
| SLCO4A1:RP11-93B14.5 | rs75880235 | chr20:61297886 | | T/C | | 0.98 | |  | |  | |  | |  | |  | |  | |  | |  | |  | |  | |  | |  | |  |
| C21orf54 | rs895438538 | chr21:34553861 | | CA/C | | 0.33 | | d/i | | -0.05 | | 7.3E-01 | | 4965 | | No | | -0.17 | | 4.0E-01 | | 1865 | | No | | 0.00 | | 2.4E-01 | | 284132 | | No |
| Diffuse opacities (quantitative) |  |  | |  | |  | |  | |  | |  | |  | |  | |  | |  | |  | |  | |  | |  | |  | |  |
| Y_RNA | rs77669438 | chr7:155959907 | | A/G | | 0.98 | |  | |  | |  | |  | |  | |  | |  | |  | |  | |  | |  | |  | |  |
| CTA-21C21.1 | rs1658697761 | chr1:88730619 | | CA/C | | 0.11 | | d/i | | 0.03 | | 5.8E-01 | | 4965 | | Yes | | 0.09 | | 1.9E-01 | | 1865 | | Yes | | -0.01 | | 6.2E-02 | |  | | No |
| Hypoplastic defects (quantitative) | | |  | |  | |  | |  | |  | |  | |  | |  | |  | |  | |  | |  | |  | |  | |  | |
| RP11-157E21.2 | rs192067416 | chr8:134990870 | | G/A | | 0.52 | | a/g | | -0.14 | | 8.4E-01 | | 1679 | | Yes | |  | |  | |  | |  | |  | |  | |  | |  |
| AC007950.1 | rs79505735 | chr15:63756376 | | C/T | | 0.46 | |  | |  | |  | |  | |  | |  | |  | |  | |  | |  | |  | |  | |  |
| RP1-167G20.1 | rs149507176 | chr5:16358142 | | C/T | | 0.52 | |  | |  | |  | |  | |  | |  | |  | |  | |  | |  | |  | |  | |  |
| ADAMTSL1 | rs115373518 | chr9:18912869 | | G/A | | 0.32 | | a/g | | 0.51 | | 3.8E-01 | | 3374 | | No | |  | |  | |  | |  | |  | |  | |  | |  |
| RAVER2 | -- | chr1:65295917 | | T/TG | | 0.52 | | d/i | | 0.12 | | 7.6E-01 | | 1232 | | Yes | | 0.29 | | 4.3E-01 | | 1364 | | Yes | | 0.01 | | 2.6E-01 | |  | | Yes |
| FHIT | rs73835238 | chr3:60260100 | | G/A | | 0.23 | |  | |  | |  | |  | |  | |  | |  | |  | |  | |  | |  | |  | |  |
| UBE2G2 | rs115617269 | chr21:46187102 | | A/G | | 0.98 | |  | |  | |  | |  | |  | |  | |  | |  | |  | |  | |  | |  | |  |
|  | rs115478160** | chr21:46257534 | |  | |  | | a/c | | 0.90 | | 3.7E-01 | | 957 | | Yes | |  | |  | |  | |  | |  | |  | |  | |  |
| MPV17L2 | rs79028494 | chr19:18296538 | | A/G | | 0.52 | | a/g | | 0.06 | | 2.6E-01 | | 18999 | | Yes | | 0.10 | | 9.9E-02 | | 13384 | | Yes | | -0.01 | | 1.9E-01 | |  | | No |
| PPAPDC3 | rs75061854 | chr9:134174252 | | T/C | | 0.52 | | t/c | | 0.47 | | 6.2E-01 | | 426 | | Yes | | 0.35 | | 7.1E-01 | | 426 | | Yes | |  | |  | |  | |  |
| CLUL1 | rs114989924 | chr18:598855 | | G/A | | 0.37 | |  | |  | |  | |  | |  | |  | |  | |  | |  | |  | |  | |  | |  |
| MAML2 | rs150489138 | chr11:96003497 | | A/G | | 0.52 | |  | |  | |  | |  | |  | |  | |  | |  | |  | |  | |  | |  | |  |
| MTHFD1L | rs11963290 | chr6:151269434 | | T/C | | 0.52 | | t/c | | 0.15 | | 1.3E-01 | | 18221 | | Yes | | 0.01 | | 9.4E-01 | | 12262 | | Yes | | 0.00 | | 7.3E-01 | |  | | Yes |
| Diffuse opacities (binary) |  |  | |  | |  | |  | |  | |  | |  | |  | |  | |  | |  | |  | |  | |  | |  | |  |
| RP1-281H8.3 | rs184135739 | chr6:149816701 | | C/T | | 0.98 | |  | |  | |  | |  | |  | |  | |  | |  | |  | |  | |  | |  | |  |

**Proxy SNP with R^2^>0.80; GLIDE children and adult study (Haworth et al. 2018; Shungin et al. 2019).
Abbreviations: LD: Linkage Disequilibrium; EA: Effect allele; OA: Other allele; P: P-value; N: sample size; chr: Chromosome; pos: Position; DC: Directional consistency

**Appendix Table 14.** Generalization of genome-wide significant loci identified in the GWAS accounting for sex (Approach 2 and 3) to GLIDE children and GLIDE adults study results.

| Locus name | rsid | Chr:Pos(hg19) | EA/OA | Beta | GLIDE | GLIDE children (Primary teeth_multiethnic) | | | | GLIDE children (Permanent teeth) | | | | GLIDE adults (DMFS and Dentures traits) | | | |
| --- | --- | --- | --- | --- | --- | --- | --- | --- | --- | --- | --- | --- | --- | --- | --- | --- | --- |
|  |  |  |  |  | **EA/OA** | **Beta** | **P** | **N** | **DC** | **Beta** | **P** | **N** | **DC** | **Beta** | **P** | **N** | **DC** |
| Developmental defects of enamel (Quantitative) | | | | |  |  |  |  |  |  |  |  |  |  |  |  |  |
| ALDH1A1** | rs141753205 | chr9:75600523 | T/C | 0.58 |  |  |  |  |  |  |  |  |  |  |  |  |  |
| RP11-384F7.1 | rs202021524 | chr3:118027270 | T/TA | 0.51 | d/i | -0.89 | 3.6E-01 | 214 | No | -0.89 | 3.6E-01 | 214 | No |  |  |  |  |
| TMPRSS13 | rs117786109 | chr11:117819775 | A/G | 0.49 | a/g |  |  |  |  | 0.07 | 1.5E-01 | 12824 | Yes | -0.01 | 1.5E-01 | 285248 | No |
| Y_RNA | rs143210152 | chr7:155957044 | G/C |  |  |  |  |  |  |  |  |  |  |  |  |  |  |
| Demarcated opacities (Quantitative) | | |  |  |  |  |  |  |  |  |  |  |  |  |  |  |  |
| *JRKL:JRKL-AS1* | rs141989840 | chr11:96225204 | A/T | 0.49 |  |  |  |  |  |  |  |  |  |  |  |  |  |
|  | rs7948994*** | chr11:96220933 |  |  | t/c | 1.23 | 9.2E-02 | 957 | No |  |  |  |  |  |  |  |  |
| *DLGAP2* | rs2906574 | chr8:1503312 | C/T | -0.53 |  |  |  |  |  |  |  |  |  |  |  |  |  |
|  | rs2906575*** | chr8:1506551 |  |  | c/g | -0.04 | 5.0E-01 | 18990 | Yes | 0.05 | 4.0E-01 | 13385 | No | 0.00 | 4.4E-01 | 285248 | Yes |
| *RP11-57H14.3*** | rs77466522 | chr10:114674436 | T/G | 0.58 | t/g | 0.11 | 4.4E-01 | 16852 | Yes | -0.02 | 8.9E-01 | 12563 | No |  |  |  |  |
| *SLC2A3* | rs61921622 | chr12:8070640 | A/G |  |  |  |  |  |  |  |  |  |  |  |  |  |  |
| Diffuse opacities (Quantitative) | | | | |  |  |  |  |  |  |  |  |  |  |  |  |  |
| *GBE1*** | rs57583981 | chr3:81739875 | T/G | 0.59 |  |  |  |  |  |  |  |  |  |  |  |  |  |
| *TENM2**^ⴕ^* | rs141872488 | chr5:167492994 | C/T | 0.58 |  |  |  |  |  |  |  |  |  |  |  |  |  |
| *RP11-384F7.1* | -- | chr3:118027270 | T/TA | 0.51 | d/i | -0.89 | 3.6E-01 | 214 | No | -0.89 | 3.6E-01 | 214 | No |  |  |  |  |
| *Y_RNA* | rs77669438 | chr7:155959907 | A/G |  |  |  |  |  |  |  |  |  |  |  |  |  |  |
| *CTA-21C21.1* | rs11440629 | chr1:88730619 | CA/C |  | d/i | 0.03 | 5.8E-01 | 4965 |  | 0.09 | 1.9E-01 | 1865 |  | -0.01 | 6.2E-02 | 284132 |  |
| Hypoplastic defects (Quantitative) | | | | |  |  |  |  |  |  |  |  |  |  |  |  |  |
| *KRTAP7-1*** | rs111896551 | chr21:32205205 | C/T | 0.59 |  |  |  |  |  |  |  |  |  |  |  |  |  |
| *CCSER1*** | rs139393106 | chr4:92616784 | T/A | 0.55 |  |  |  |  |  |  |  |  |  |  |  |  |  |
|  | rs182744873*** | chr4:92577649 |  |  | t/c | -0.38 | 7.3E-01 | 235 | No | -0.16 | 8.9E-01 | 234 | No |  |  |  |  |
| *PLEKHD1* | rs147589787 | chr14:69956684 | C/A | 0.56 | a/c | -0.29 | 3.5E-01 | 5208 | Yes | -0.03 | 9.8E-01 | 396 | Yes |  |  |  |  |
| *MAST4* | rs1753671813 | chr5:66314987 | A/AT | 0.55 | d/i | 0.29 | 6.5E-01 | 957 | Yes |  |  |  |  |  |  |  |  |
| *NF1* | rs147836184 | chr17:29427876 | A/G | 0.58 |  |  |  |  |  |  |  |  |  |  |  |  |  |
| *CCDC18* | rs187188755 | chr1:93677880 | T/A | 0.55 | a/t | 0.02 | 9.2E-01 | 12671 | No | 0.09 | 6.6E-01 | 10979 | No |  |  |  |  |
| *TENM2* | rs62383285 | chr5:166907015 | A/G | 0.56 | a/g | 0.04 | 7.2E-01 | 14384 | Yes | -0.23 | 2.6E-02 | 13033 | No | 0.01 | 8.7E-02 | 285247 | Yes |
| *AC007950.1* | rs79505735 | chr15:63756376 | C/T |  |  |  |  |  |  |  |  |  |  |  |  |  |  |
| *RP1-167G20.1* | rs149507176 | chr5:16358142 | C/T |  |  |  |  |  |  |  |  |  |  |  |  |  |  |
| *RP11-157E21.2* | rs192067416 | chr8:134990870 | G/A |  | a/g | -0.14 | 8.4E-01 | 1679 |  |  |  |  |  |  |  |  |  |
| Developmental defects of enamel (Binary) | | | | |  |  |  |  |  |  |  |  |  |  |  |  |  |
| SLC19A1 | rs62214342 | chr21:46998930 | A/G | 0.34 | a/g | 0.07 | 2.3E-02 | 19002 | Yes | 0.02 | 6.2E-01 | 13386 | Yes | 0.00 | 1.1E-01 | 285247 | Yes |
| snoU13 | rs7228955 | chr18:19840997 | G/C |  | c/g | -0.04 | 2.4E-01 | 18993 |  | 0.01 | 8.0E-01 | 13382 |  | -0.01 | 8.2E-03 | 285248 |  |
| Demarcated opacities (Binary) | | | | |  |  |  |  |  |  |  |  |  |  |  |  |  |
| *CTD-2644I21.1*** | rs2776544 | chr14:101054029 | G/T | 0.34 | t/g | 0.02 | 5.2E-01 | 18952 | No | 0.06 | 5.4E-02 | 13371 | No | 0.00 | 2.6E-01 | 285246 | No |
| *RP11-203H2.1* | rs17143504 | chr6:8292791 | T/G | -0.44 |  |  |  |  |  | 0.03 | 4.8E-01 | 7999 |  |  |  |  |  |
|  | rs62398162*** | chr6:8293455 |  |  | a/c | -0.02 | 6.6E-01 | 19003 | Yes | 0.02 | 5.6E-01 | 13386 | No | 0.00 | 9.1E-01 | 285249 | No |
| Diffuse opacities (Binary) | | | | |  |  |  |  |  |  |  |  |  |  |  |  |  |
| *LINC00670* | rs11078084 | chr17:12513995 | T/C | -0.29 | t/c | 0.02 | 4.0E-01 | 19002 | No | 0.03 | 3.5E-01 | 13386 | No | 0.00 | 7.4E-01 | 285247 | Yes |

**Proxy SNP with R^2^>0.80 (LD); GLIDE children and adult study (Haworth et al. 2018; Shungin et al. 2019). ** SNP is also genome-wide statistically significant for the joint test (main genetic effect + gene x environment effect), ⴕ SNP is also genome-wide statistically significant for the test of difference (i.e. interaction effect)Abbreviations: LD: Linkage Disequilibrium; EA: Effect allele; OA: Other allele; P: P-value; N: sample size; chr: Chromosome; pos: Position; DC: Directional consistency

**Appendix table 15.** Cross-phenotype p-value landscape of genome-wide significant loci from GWAS of developmental defects of enamel traits

| **rsid** | **Locus name** | **dde** | **dde_count** | **dema** | **dema_count** | **diff** | **diff_count** | **hypo** | **hypo_count** |
| --- | --- | --- | --- | --- | --- | --- | --- | --- | --- |
| rs567243779 | *DEK* | 4.43E-02 | **8.09E-09** | 9.38E-03 | 4.90E-05 | 2.09E-04 | 9.78E-06 | 3.98E-01 | 5.61E-02 |
| rs116989668 | *HBS1L* | 8.20E-02 | **4.02E-08** | 1.85E-01 | 1.28E-05 | 3.64E-01 | 9.44E-03 | 6.82E-02 | 5.17E-03 |
| rs150075871 | *RP4-724E13.2* | 1.31E-02 | **1.98E-08** | 3.30E-02 | 4.03E-05 | 1.47E-03 | 9.90E-02 | 5.58E-02 | 2.40E-06 |
| rs75880235 | *SLCO4A1:RP11-93B14.5* | 5.19E-01 | 9.48E-06 | 1.21E-01 | **1.89E-08** | 3.98E-01 | 1.80E-01 | 7.30E-01 | 6.61E-01 |
| rs895438538 | *C21orf54* | 6.50E-03 | 7.63E-06 | 2.89E-04 | **4.20E-08** | 2.24E-01 | 1.80E-01 | 6.10E-02 | 9.26E-02 |
| rs1658697761 | *CTA-21C21.1* | 7.48E-01 | 1.75E-06 | 6.75E-02 | 2.21E-02 | 2.27E-03 | **2.02E-09** | 5.40E-02 | 4.83E-01 |
| rs184135739 | *RP1-281H8.3* | 1.07E-03 | 5.47E-04 | 2.77E-01 | 9.68E-02 | **2.95E-09** | 2.35E-04 | 4.09E-01 | 6.09E-01 |
| rs143210152 | *Y_RNA* | 1.18E-03 | **4.45E-09** | 1.02E-02 | 4.69E-03 | 7.73E-04 | **5.19E-10** | 2.63E-03 | 2.91E-03 |
| rs77669438 | *Y_RNA* | 7.16E-03 | **3.84E-08** | 3.66E-02 | 4.78E-02 | 9.38E-04 | **9.12E-11** | 2.08E-03 | 2.00E-03 |
| chr1:64830234 | *RAVER2* | 3.67E-01 | 5.94E-01 | 8.62E-02 | 2.47E-01 | 4.03E-01 | 6.57E-01 | 3.43E-02 | **7.61E-09** |
| rs73835238 | *FHIT* | 5.92E-01 | 1.99E-02 | 5.39E-01 | 1.79E-01 | 8.59E-01 | 7.36E-01 | 4.14E-03 | **9.11E-09** |
| rs149507176 | *RP1-167G20.1* | 1.40E-01 | 5.45E-02 | 7.69E-01 | 6.71E-01 | 6.32E-01 | 9.13E-01 | 4.45E-06 | **1.46E-09** |
| rs11963290 | *MTHFD1L* | 1.02E-01 | 1.60E-05 | 4.39E-01 | 2.38E-01 | 4.22E-03 | 4.20E-04 | 3.72E-03 | **4.80E-08** |
| rs192067416 | *RP11-157E21.2* | 3.01E-01 | 3.31E-03 | 4.97E-01 | 2.28E-01 | 7.78E-01 | 5.36E-01 | 7.94E-05 | **2.43E-10** |
| rs115373518 | *ADAMTSL1* | 2.11E-02 | 5.42E-03 | 1.24E-01 | 5.24E-01 | 1.07E-01 | 1.70E-01 | 1.69E-04 | **7.46E-09** |
| rs75061854 | *PPAPDC3* | 6.12E-01 | 3.72E-01 | 8.85E-01 | 4.98E-01 | 8.58E-01 | 4.51E-01 | 1.09E-04 | **2.93E-08** |
| rs150489138 | *MAML2* | 4.67E-01 | 4.91E-02 | 6.39E-01 | 4.00E-01 | 8.97E-01 | 8.67E-01 | 1.21E-02 | **4.25E-08** |
| rs79505735 | *AC007950.1* | 1.35E-01 | 5.16E-03 | 1.62E-01 | 3.66E-01 | 4.41E-01 | 3.79E-01 | 2.79E-03 | **5.14E-10** |
| rs114989924 | *CLUL1* | 3.51E-01 | 8.85E-04 | 6.67E-01 | 5.28E-01 | 2.00E-01 | 1.71E-03 | 2.63E-05 | **3.55E-08** |
| rs79028494 | *MPV17L2* | 9.42E-01 | 6.01E-01 | 3.29E-01 | 5.35E-02 | 7.27E-01 | 4.86E-01 | 5.28E-03 | **2.88E-08** |
| rs115617269 | *UBE2G2* | 6.79E-02 | 5.29E-02 | 5.87E-01 | 4.96E-01 | 6.05E-01 | 9.84E-02 | 2.87E-03 | **2.00E-08** |
| rs202021524 | *RP11-384F7.1* | 8.21E-03 | 7.00E-05 | 3.78E-02 | 5.12E-02 | 1.56E-01 | 1.26E-03 | 4.97E-04 | 7.78E-03 |
| rs141753205 | *ALDH1A1* | 1.65E-03 | 1.25E-05 | 1.65E-03 | 1.23E-03 | 7.86E-02 | 5.92E-03 | 3.12E-01 | 4.53E-02 |
| rs117786109 | *TMPRSS13* | 2.68E-02 | 1.18E-05 | 1.45E-02 | 9.44E-03 | 1.41E-02 | 9.28E-05 | 6.86E-01 | 3.57E-01 |
| rs7228955 | *snoU13* | 3.02E-01 | 4.85E-01 | 8.67E-01 | 8.06E-02 | 9.59E-01 | 6.53E-01 | 4.98E-01 | 6.47E-01 |
| rs62214342 | *SLC19A1* | 5.55E-03 | 1.16E-01 | 7.21E-02 | 9.41E-02 | 7.24E-02 | 6.06E-01 | 6.44E-01 | 5.94E-01 |
| rs17143504 | *RP11-203H2.1* | 1.65E-05 | 1.84E-04 | 1.01E-05 | 1.29E-04 | 1.52E-03 | 4.55E-02 | 1.23E-01 | 8.10E-01 |
| rs2906574 | *DLGAP2* | 5.38E-02 | 1.36E-01 | 4.27E-02 | 5.75E-03 | 6.18E-01 | 4.24E-01 | 7.06E-01 | 4.14E-01 |
| rs77466522 | *RP11-57H14.3* | 1.40E-02 | 1.48E-03 | 3.47E-01 | 4.75E-04 | 6.05E-01 | 6.96E-01 | 1.85E-04 | 3.37E-03 |
| rs141989840 | *JRKL:JRKL-AS1* | 9.69E-03 | 7.28E-05 | 1.15E-03 | 1.21E-06 | 1.62E-01 | 1.91E-01 | 2.84E-01 | 5.97E-01 |
| rs61921622 | *SLC2A3* | 6.44E-01 | 5.40E-01 | 8.36E-01 | 1.75E-01 | 2.38E-01 | 8.07E-01 | 3.96E-01 | 9.15E-01 |
| rs2776544 | *CTD-2644I21.1* | 8.74E-02 | 1.57E-01 | 1.27E-05 | 9.37E-03 | 1.29E-01 | 5.52E-01 | 8.85E-01 | 3.99E-01 |
| rs57583981 | *GBE1* | 9.58E-03 | 2.85E-03 | 2.30E-01 | 7.55E-01 | 8.18E-02 | 4.64E-07 | 2.45E-01 | 3.45E-01 |
| rs11078084 | *LINC00670* | 1.11E-03 | 2.11E-03 | 7.38E-02 | 2.20E-02 | 1.15E-05 | 7.72E-03 | 1.00E+00 | 5.14E-01 |
| rs141872488 | *TENM2* | 1.27E-01 | 1.27E-01 | 7.30E-01 | 9.46E-01 | 2.86E-01 | 3.48E-02 | 4.60E-01 | 2.14E-01 |
| rs62383285 | *TENM2* | 4.36E-01 | 9.18E-02 | 5.25E-01 | 7.73E-01 | 1.11E-01 | 2.87E-01 | 3.57E-05 | 4.24E-07 |
| rs187188755 | *CCDC18* | 8.65E-02 | 7.59E-01 | 2.48E-01 | 9.99E-01 | 1.59E-01 | 2.25E-01 | 4.78E-01 | 8.19E-04 |
| rs139393106 | *CCSER1* | 9.19E-01 | 3.14E-02 | 8.03E-01 | 4.85E-01 | 3.08E-01 | 5.43E-01 | 8.32E-03 | 3.34E-06 |
| rs1753671813 | *MAST4* | 3.83E-01 | 3.74E-01 | 2.12E-02 | 1.64E-01 | 3.98E-01 | 2.69E-01 | 2.62E-02 | 6.46E-05 |
| rs147589787 | *PLEKHD1* | 7.05E-01 | 4.86E-02 | 7.61E-01 | 7.65E-01 | 7.26E-01 | 2.82E-01 | 9.16E-03 | 8.48E-07 |
| rs147836184 | *NF1* | 6.28E-01 | 2.34E-01 | 5.61E-01 | 9.09E-01 | 7.46E-01 | 8.76E-01 | 5.31E-01 | 3.05E-04 |
| rs111896551 | *KRTAP7-1* | 1.06E-01 | 8.73E-02 | 6.52E-01 | 4.58E-01 | 9.88E-01 | 7.77E-01 | 2.20E-02 | 2.56E-06 |

**p-value**

Not significant (p ≥ 0.05)

p < 0.05

Suggestive (p < 10⁻⁵)

Genome-wide significant loci in the main discovery analysis

Genome-wide significant loci in the sex-stratified analysis

dde: combined developmental defects of enamel

dema: demarcated opacities

diff: diffuse opacities

hypo: hypoplastic defects

count: quantitative trait denoting the number of affected teeth

**Appendix Table 16.** Association of the *SCUBE* locus, rs13058467 (lowest p-value in Kühnisch et al. 2014) in the present study’s results for DDE.

| **Trait** | **EA** | **Freq (%)** | **OR** | **CI** | **P-value** |
| --- | --- | --- | --- | --- | --- |
| MIH | C | 10.40 | 4.38 | 2.48-0.4 | 3.70E-07 |

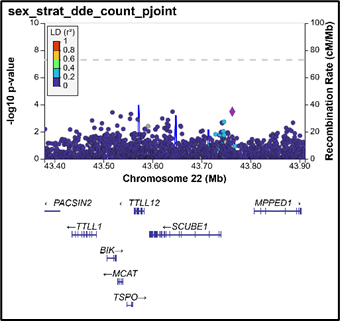

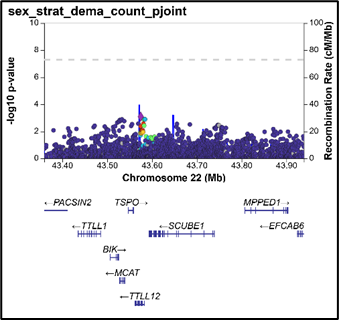

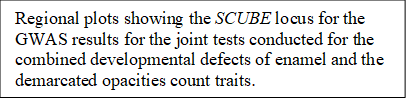

| **QUANTITATIVE** | **BETA.female** | **p.value.female** | **BETA.male** | **p.value.male** | **P.diff** | **P.joint** | **Concordant Direction** |
| --- | --- | --- | --- | --- | --- | --- | --- |
| dde_count | 0.11 | **3.23E-02** | -0.09 | 8.52E-02 | 5.86E-03 | **2.30E-02** | Y |
| dema_count | 0.15 | **2.69E-03** | -0.04 | 4.66E-01 | 7.53E-03 | **8.50E-03** | Y |
| diff_count | -0.04 | 3.85E-01 | -0.05 | 3.39E-01 | 9.55E-01 | 4.34E-01 |  |
| hypo_count | 0.08 | 1.22E-01 | -0.12 | **1.81E-02** | 5.61E-03 | **1.85E-02** | N |
| **BINARY** | **BETA.female** | **p.value.female** | **BETA.male** | **p.value.male** | **P.diff** | **P.joint** |  |
| hypo | 0.14 | 2.56E-01 | -0.31 | **1.06E-02** | 8.71E-03 | **1.99E-02** | N |
| dde | 0.21 | 5.63E-02 | 0.02 | 8.70E-01 | 2.13E-01 | 1.60E-01 |  |
| dema | 0.17 | 9.64E-02 | 0.05 | 6.23E-01 | 3.99E-01 | 2.22E-01 |  |
| diff | 0.13 | 2.03E-01 | 0.05 | 5.99E-01 | 5.81E-01 | 3.87E-01 |  |

*chr22:43183043; EAF=0.07, EA/OA=C/T

**Abbreviations:** MIH: Molar-incisor hypomineralization, CHR: chromosome, SNP: Single nucleotide polymorphism, EA: Effect Allele, EAF: Effect allele frequency, OR: Odds ratio, CI: Confidence interval; dde: developmental defects of enamel; dema: demarcated opacities, diff: diffuse opacities, hypo: hypoplastic defects, effN: effective sample size, ctrl: Control.
