## Supplementary material for "Genomic basis of developmental defects of enamel and sex-specific effects": Main Figures

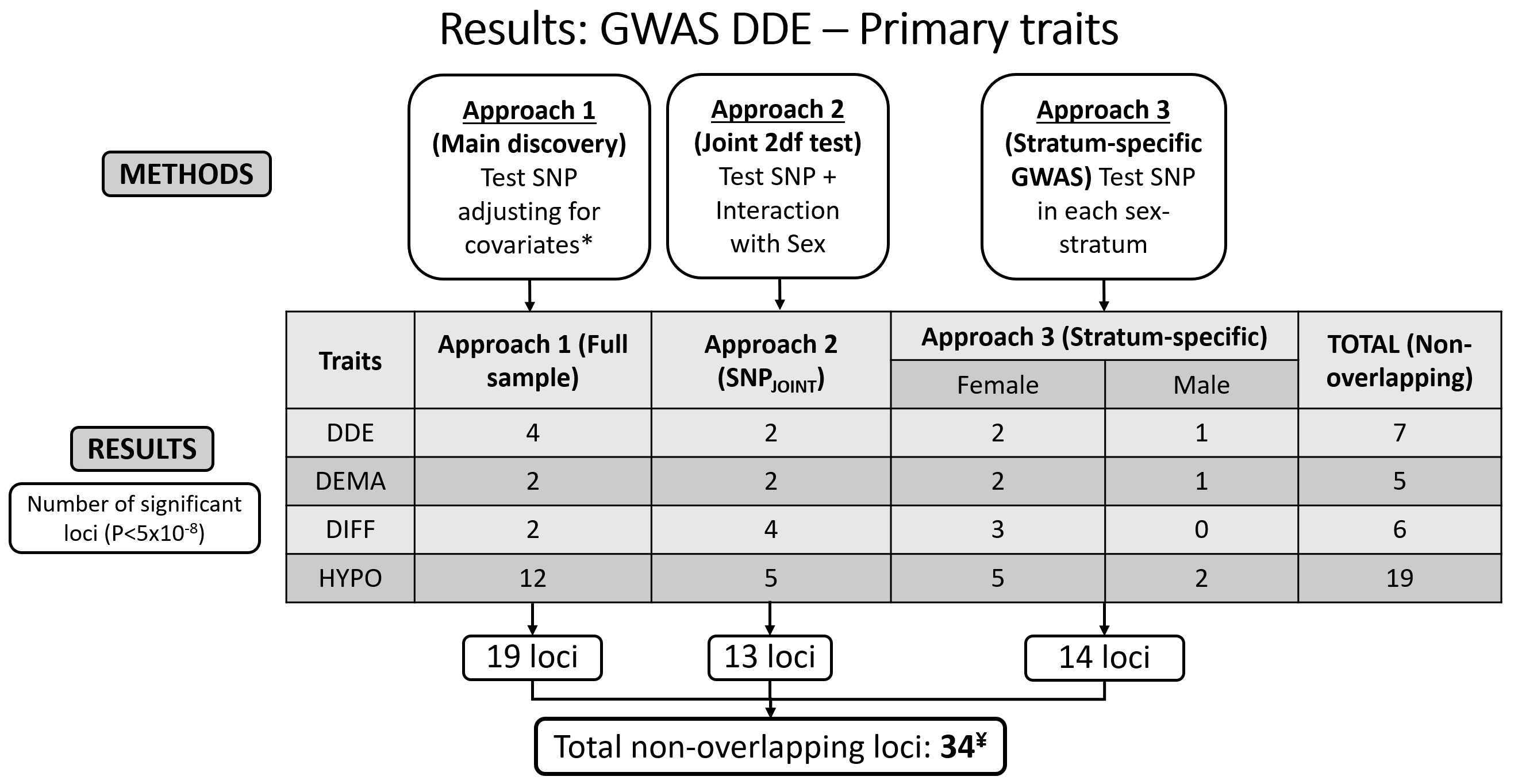

Figure 1: Summary of study design and results for primary count traits. Approach 1 conducted a genome-wide association study (GWAS) in the entire sample. Approach 2 conducted a joint 2-degree-of-freedom test to test the main and interaction effects jointly accounting for sex. Approach 3 conducted sex-specific GWAS of the developmental defects of enamel primary quantitative traits. The approaches have been detailed in the Methods section. The total number of loci is less than the sum of the boxes owing to the identification of the same loci in different traits or analyses. *Covariates include age, sex, race/ethnicity, and the first 8 principal components of ancestry. ¥ Analysis of secondary binary traits identified further 5 independent genomic risk loci. Abbreviations: DDE: Developmental Defects of Enamel, DEMA: Demarcated Opacities, DIFF: Diffuse Opacities, HYPO: Hypoplastic defects, SNP: Single Nucleotide Polymorphism, 2df: 2-degree-of-freedom, GWAS: Genome-wide association study, P: p-value.

| 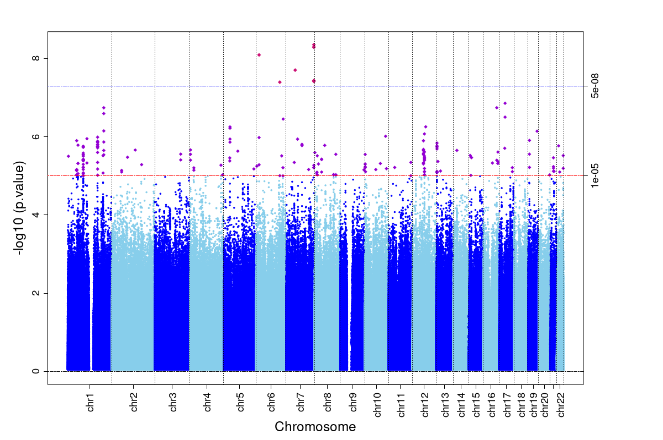  (A) | 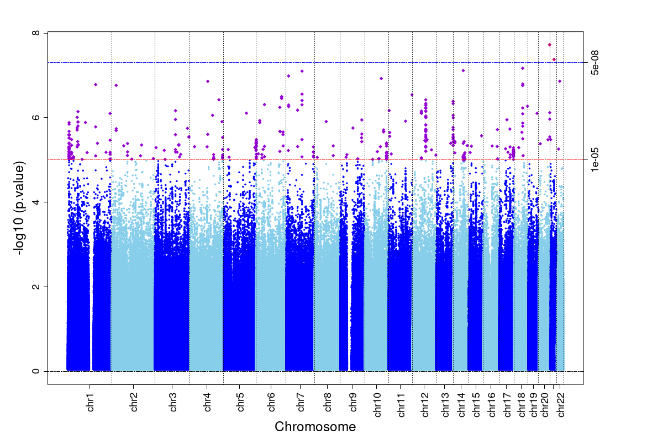  (B) |
| --- | --- |
| 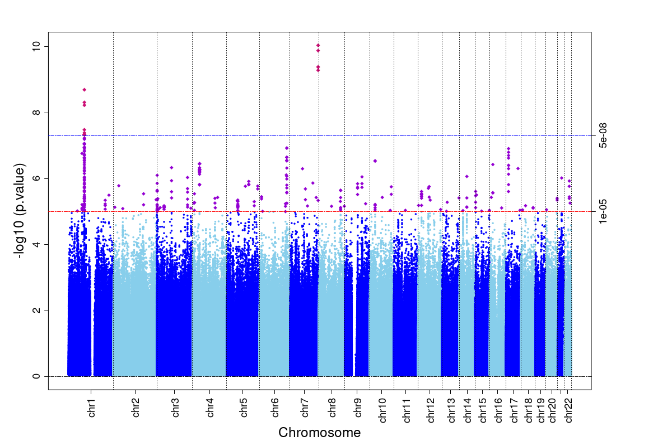  (C) | 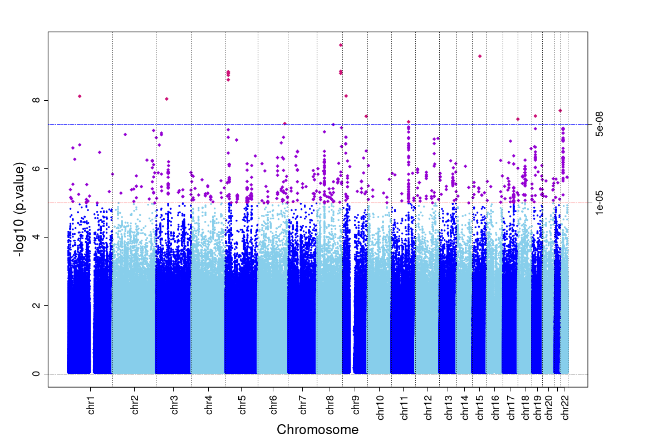  (D) |

Figure 2: Manhattan plots for the primary quantitative traits with genome-wide statistically significant loci (Approach 1). A) DDE, B) Demarcated opacities, C) Diffuse Opacities, D) Hypoplastic defects

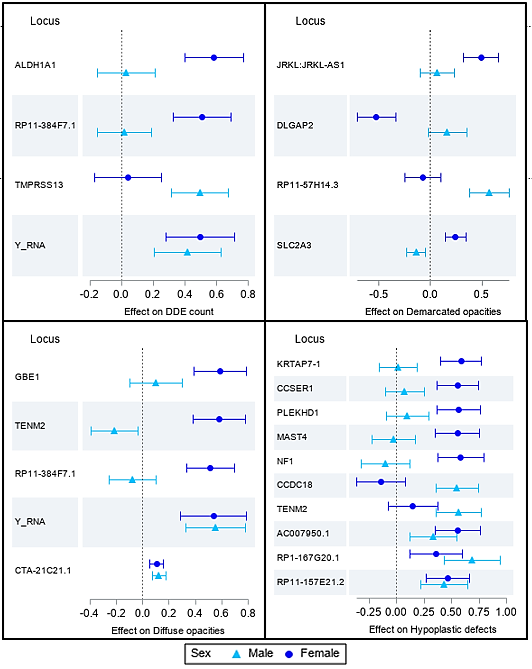

D

C

A

B

Figure 3: Forest plots demonstrating heterogeneity of genetic effect in different sexes for the 4 primary traits. (A) Developmental defects of enamel count, (B) Demarcated opacities, (C) Diffuse opacities, (D) Hypoplastic defects. The triangles and circles represent the effect estimates of association with the trait and the error bars represent the corresponding 95% confidence intervals.
